# Consistency, completeness and external validity of ethnicity recording in NHS primary care records: a cohort study in 25 million patients’ records at source using OpenSAFELY

**DOI:** 10.1101/2023.11.21.23298690

**Authors:** The OpenSAFELY Collaborative, Colm D Andrews, Rohini Mathur, Jon Massey, Robin Park, Lisa Hopcroft, Helen J Curtis, Amir Mehrkar, Seb Bacon, George Hickman, Rebecca Smith, David Evans, Tom Ward, Simon Davy, Peter Inglesby, Iain Dillingham, Steven Maude, Thomas O’Dwyer, Ben Butler-Cole, Lucy Bridges, Chris Bates, John Parry, Frank Hester, Sam Harper, Jonathan Cockburn, Ben Goldacre, Brian MacKenna, Laurie Tomlinson, Alex J Walker, William J Hulme

**Affiliations:** Bennett Institute for Applied Data Science, Nuffield Department of Primary Care Health Sciences, Oxford University, Oxford, OX2 6GG; London School of Hygiene and Tropical Medicine, Keppel Street, London WC1E 7HT; TPP, TPP House, 129 Low Lane, Horsforth, Leeds, LS18 5PX; Wolfson Institute for Population Health, Queen Mary, University of London, London, E1 2AT

## Abstract

**Background:** Ethnicity is known to be an important correlate of health outcomes, particularly during the COVID-19 pandemic, where some ethnic groups were shown to be at higher risk of infection and adverse outcomes. The recording of patients’ ethnic groups in primary care can support research and efforts to achieve equity in service provision and outcomes; however the coding of ethnicity is known to present complex challenges. We therefore set out to describe ethnicity coding in detail with a view to supporting the use of this data in a wide range of settings, as part of wider efforts to robustly describe and define methods of using administrative data.

**Methods:** We describe the completeness and consistency of primary care ethnicity recording in the OpenSAFELY-TPP database, containing linked primary care and hospital records in >25 million patients in England. We also compared the ethnic breakdown in OpenSAFELY-TPP with that of the 2021 UK census.

**Results:** 78.2% of patients registered in OpenSAFELY-TPP on 1 January 2022 had their ethnicity recorded in primary care records, rising to 92.5% when supplemented with hospital data. The completeness of ethnicity recording was higher for women than for men. The rate of primary care ethnicity recording ranged from 77% in the South East of England to 82.2% in the West Midlands. Ethnicity recording rates were higher in patients with chronic or other serious health conditions. For each of the five broad ethnicity groups, primary care recorded ethnicity was within 2.9 percentage points of the population rate as recorded in the 2021 Census for England as a whole. For patients with multiple ethnicity records, 98.7% of the latest recorded ethnicities matched the most frequently coded ethnicity. Patients whose latest recorded ethnicity was categorised as Other were most likely to have a discordant ethnicity recording (32.2%).

**Conclusions:** Primary care ethnicity data in OpenSAFELY is present for over three quarters of all patients, and combined with data from other sources can achieve a high level of completeness. The overall distribution of ethnicities across all English OpenSAFELY-TPP practices was similar to the 2021 Census, with some regional variation. This report identifies the best available codelist for use in OpenSAFELY and similar electronic health record data.

## Background

Ethnicity is known to be an important determinant of health inequalities, particularly during the COVID-19 outbreak where a complex interplay of social and biological factors resulted in increased exposure, reduced protection, and increased severity of illness in particular ethnic groups^1,2^. The UK has a diverse ethnic population (The 2021 ONS Census estimated 9.6% Asian, 4.2% Black, 3.0% Mixed, 81.0% White, 2.2% Other^3^), which can make health research conducted in the UK generalisable to countries. Complete and consistent recording of patients’ ethnic group in primary care can support efforts to achieve equity in service provision and reduces bias in research^4,5^. Ethnicity recording for new patients registering with general practice across the UK has improved following Quality and Outcomes Framework (QOF) financial incentivization between 2006/07 and 2011/12^6,7^. As a result, ethnicity is now being captured for the majority of the population in routine electronic healthcare records, and is comparable to the general population^6^. The uptake and utilisation of healthcare services still varies across ethnic groups and the recently established NHS Race and Health Observatory have led calls for a dedicated drive by NHS England and NHS Digital to emphasise the importance of collecting and reporting ethnicity data^8^.

OpenSAFELY is a secure health analytics platform created by our team on behalf of NHS England. OpenSAFELY provides a secure software interface allowing analysis of pseudonymised primary care patient records from England in near real-time within highly secure data environments.

In primary care data, patient ethnicity is recorded via clinical codes, similar to how any other clinical condition or event is recorded. In OpenSAFELY-TPP, both Clinical Terms Version 3 (CTV3 (Read)) codes and Systematised Nomenclature of Medicine Clinical Terms (SNOMED CT) codes are used. SNOMED CT is an NHS standard, widely used across England.

Ethnicity is also recorded in secondary care, when patients attend emergency care, inpatient or outpatient services, independently of ethnicity in the primary care record. This is available via NHS England’s Secondary Uses Service (SUS)^9^. It is common practice in OpenSAFELY to supplement primary care ethnicity, where missing, with ethnicity data from SUS^10,11^.

In this paper, we study the completeness, consistency and representativeness of routinely collected ethnicity data in primary care.

## Methods

### Study design

Retrospective cohort study across 25m patients registered with English general practices in OpenSAFELY-TPP.

### Data Sources

This study uses data from the OpenSAFELY-TPP database, covering around 40% of the English population. The database includes primary care records of patients in practices using the TPP SystmOne patient information system, and is linked to other NHS data sources, including in-patient hospital records from NHS England’s Secondary Use Service (SUS), where ethnicity is also recorded independently of ethnicity in the primary care record.

All data were linked, stored and analysed securely within the OpenSAFELY platform https://opensafely.org/. **Data include pseudonymized data such as coded diagnoses, medications and physiological parameters. No free text data are included**. All code is shared openly for review and re-use under MIT open licence (<u>opensafely/ethnicity-short-data-report at notebook</u>). Detailed pseudonymised patient data is potentially re-identifiable and therefore not shared. We rapidly delivered the OpenSAFELY data analysis platform without prior funding to deliver timely analyses on urgent research questions in the context of the global COVID-19 health emergency: now that the platform is established we are developing a formal process for external users to request access in collaboration with NHS England; details of this process are available at OpenSAFELY.org.

### Study population

Patients were included in the study if they were registered at an English general practice using TPP on 1 January 2022.

### Ethnicity ascertainment

In primary care data, there is no categorical “ethnicity” variable to record this information. Rather, ethnicity is recorded using clinical codes - entered by a clinician or administrator with a location and date - like any other clinical or administrative event, with specific codes relating to each ethnic group^12–14^. This means ethnicity can be recorded by the practice in multiple, potentially conflicting, ways over time.

We created a new codelist, SNOMED:2022^13^ by identifying relevant ethnicity SNOMED CT codes and ensuring completeness by comparing the codelist to: another OpenSAFELY created codelist (CTV3:2020)^13^; a combined ethnicity codelist from SARS-CoV2 COVID19 Vaccination Uptake Reporting Codes published by Primary Care Information Services (PRIMIS)^12,15^; and a codelist from General Practice Extraction Service (GPES) Data for Pandemic Planning and Research (GDPPR)^16^. Codes which relate to religion rather than ethnicity (e.g. “Muslim - ethnic category 2001 census”) and codes which do not specify a specific ethnicity (e.g. “Ethnic group not recorded”) were excluded. In total 258 relevant ethnicity codes were identified. We then created a codelist categorisation based on the 2001 UK Census categories, which are the NHS standard for ethnicity^17^, and cross referenced it against the CTV3, PRIMIS and GDPPR codelists. The ‘Gypsy or Irish Traveller’ and ‘Arab’ groups were not specifically listed in 2001 however we categorised them as ‘White’ and ‘Other’ respectively as per the 2011 Census grouping^18^)

The codelist categorisation consists of two ethnicity groupings based on the 2001 census (Box 1): All analyses used the 5-group categorisation unless otherwise stated.

##### Box 1: 2001 ONS Census ethnicity groupings

5-level group:

- Asian or Asian British
- Black or Black British,
- Mixed,
- White
- Chinese or other ethnic group

16-level group:

- Indian
- Pakistani
- Bangladeshi
- Any other Asian background
- Caribbean, African
- Any other Black background
- White and Black
- Caribbean
- White and Black African
- White and Asian
- Any other Mixed background
- British
- Irish
- Any other White background
- Chinese
- Any other

If a SNOMED:2022 ethnicity code appeared in the primary care record on multiple dates, the latest entry was used unless otherwise stated.

In OpenSAFELY the function <u>with_ethnicity_from_sus</u> combines SUS ethnicity data from admitted patient care statistics (APCS), Emergency Care (EC) and outpatient attendance (OPA) and selects the most frequently used ethnicity code for each patient. In hospital records from SUS, recorded ethnicity is categorised as one of the 16 categories on the 2001 UK census. This accords with the 16-level grouping described above.

### Subgroups

We looked at the completeness of ethnicity coding in the whole population and across each of the following demographic and clinical subgroups:

- Age: Patient age was calculated as of 1 January 2022 and grouped into 5 year bands, to match the ONS age bands.
- Sex: We used categories “male” and “female”, matching the ONS recorded categories; patients with any other/unknown sex were excluded.
- Deprivation: Overall deprivation was measured by the 2019 Index of Multiple Deprivation (IMD)^19^ derived from the patient’s postcode at lower super output area level. IMD was divided by quintile, with 1 representing the most deprived areas and 5 representing least deprived areas. Where a patient’s postcode cannot be determined the IMD is recorded as unknown.
- Region: Region was defined as the Nomenclature of Territorial Units for Statistics (NUTS 1) region derived from the patient’s practice postcode.

As ethnicity recording would be expected to be lower in patients with fewer clinical interactions, completeness was also compared in the clinical subgroups of dementia, diabetes, hypertension and learning disability. Clinical subgroups were defined as the presence or absence of relevant SNOMED CT codes in the GP records for dementia^20^, diabetes^21^, hypertension^22^, and learning disabilities^23^ as of 1 January 2022.

### Statistical methods

#### Completeness and distribution of ethnicity recording

The proportion of patients with either (i) primary care ethnicity recorded (that is, the presence of any code in the SNOMED:2022 codelist in the patient record) or (ii) primary care ethnicity supplemented, where missing, with ethnicity data from secondary care^24^ was calculated. Completeness was reported overall and within clinical and demographic subgroups.

Amongst those patients where ethnicity was recorded, the proportion of patients within each of the 5 and 16 ethnicity groups was calculated, within each clinical and demographic subgroup.

#### Consistency of ethnicity recording within patients over time

Discrepancies may arise due to errors while entering the data or if a patient self-reports a different ethnic group from their previously recorded ethnic group. We calculated the proportion of patients with any ethnicity recorded which did not match their ‘latest’ recorded grouped ethnicity for each of the five ethnic groups.

We also calculated the proportion of patients whose latest recorded ethnicity did not match their most frequently recorded ethnicity for each of the five ethnic groups.

#### Consistency of ethnicity recording across data sources (primary care versus secondary care)

We calculated the proportion of patients whose latest recorded ethnicity in primary care matched their ethnicity as recorded in secondary care for each of the five ethnic groups.

#### External validation against the 2021 UK census population

The UK Census collects individual and household-level demographic data every 10 years for the whole UK population. Data on ethnicity were obtained from the 2021 UK Census for England. The most recent census across the UK was undertaken on 27 March 2021. Ethnic breakdowns for the population of England were obtained via NOMIS^25^.

The ethnic breakdown of the census population was compared with our OpenSAFELY-TPP population. In the 2021 UK Census the Chinese ethnic group was included in the Asian ethnic group whereas in the 2001 census it was included in the Other ethnic group^26^. In order to provide a suitable comparison with primary care data, we regrouped the 2021 census data as per the 2001 groups. As an additional analysis we also compared the primary care data with the census data using the 2021 census categories.

#### Software and Reproducibility

Data management was performed using Python 3.8, with analysis carried out using Python and R. Code for data management and analysis, as well as codelists are archived online https://github.com/opensafely/ethnicity-short-data-report/.

#### Patient and Public Involvement

This analysis relies on the use of large volumes of patient data. Ensuring patient, professional, and public trust is therefore of critical importance. Maintaining trust requires being transparent about the way OpenSAFELY works, and ensuring patient and public voices are represented in the design and use of the platform. Between February and July 2022 we ran a six month pilot of Patient and Public Involvement and Engagement activity designed to be aligned with the principles set out in the Consensus Statement on Public Involvement and Engagement with Data-Intensive Health Research^27^. Our engagement focused on the broader OpenSAFELY platform and comprised three sets of activities: explain and engage, involve and iterate and participate and promote. To engage and explain, we have developed a public website at opensafely.org that provides a detailed description of the OpenSAFELY platform in language suitable for a lay audience and are co-developing an accompanying explainer video. To involve and iterate we have created the OpenSAFELY ‘Digital Critical Friends’ Group; comprised of approximately 12 members representative in terms of ethnicity, gender, and educational background, this group has met every 2 weeks to engage with and review the OpenSAFELY website, governance process, principles for researchers and FAQs. To participate and promote, we are conducting a systematic review of the key enablers of public trust in data-intensive research and have participated in the stakeholder group overseeing NHS England’s ‘data stewardship public dialogue’.

## Results

### Completeness of ethnicity data

19,618,135 of the 25,102,210 patients (78.2%) registered in OpenSAFELY-TPP on 1 January 2022 had a recorded ethnicity, rising to 92.5% when supplemented with secondary care data (Figure 1, Extended Data Table 1).

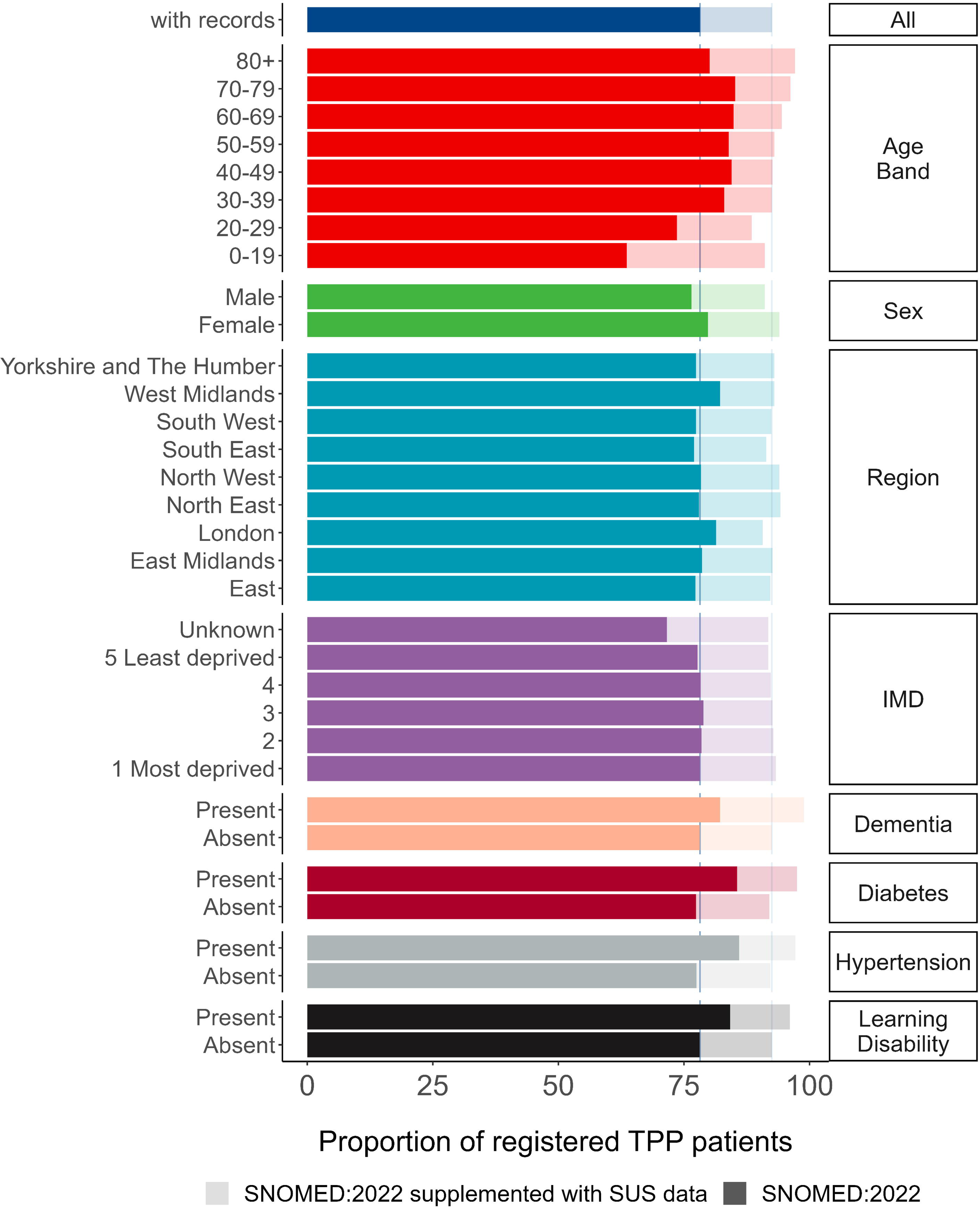

Primary care ethnicity recording completeness was lowest for patients aged over 80 years (80.1%) and under 30, whereas ethnicity recording was highest in those over 80 when supplemented with secondary care data (97.1%). Women had a higher proportion of recorded ethnicities than men (79.8% and 76.5% respectively, 94% and 91.1% when supplemented with secondary care data). The completeness of primary care ethnicity recording ranged from 77% in the South East of England to 82.2% in the West Midlands. IMD was within 1.2 percentage points for known values (77.7% in the least deprived group 5 to 78.9% in group 3) and was lowest for the unknown group (71.6%). Primary care ethnicity recording was at least 4 percentage points higher in all of the clinical subgroups compared to the general population.

### Distribution of ethnicity

Using ethnicity recorded in primary care only, 6.8% of the population were recorded as Asian, 2.3% Black, 1.5% Mixed, 65.6% White, 1.9% Other, and ethnicity was not recorded for 21.8%. When supplementing with hospital-recorded ethnicity data, corresponding percentages were 7.8% Asian, 2.6% Black, 1.9% Mixed, 77.9% White, 2.3% Other, and 7.5% not recorded, representing a percentage point increase ranging from 0.3% in the Black group to 12.3% in the White group.

Older patients tended to have a higher rate of recorded White ethnicity (e.g. 76.3% in the 80+ group vs 50.0% in the 0-19 group), whereas younger patients had a higher rate of recording for Asian, Black, Mixed and Other groups. The higher proportion of women with recorded ethnicity was reversed in the Asian group where men (7.0% and 8.0% with secondary care data) had a higher proportion of recording than women (6.6% and 7.6% with secondary care data). The proportion of ethnicity reporting was lower for patients with dementia, hypertension or learning disabilities in every ethnic group other than White (Figure 2 / Extended Data Table 2). The breakdown by 16 group ethnicity is shown in Extended Data Table 3.

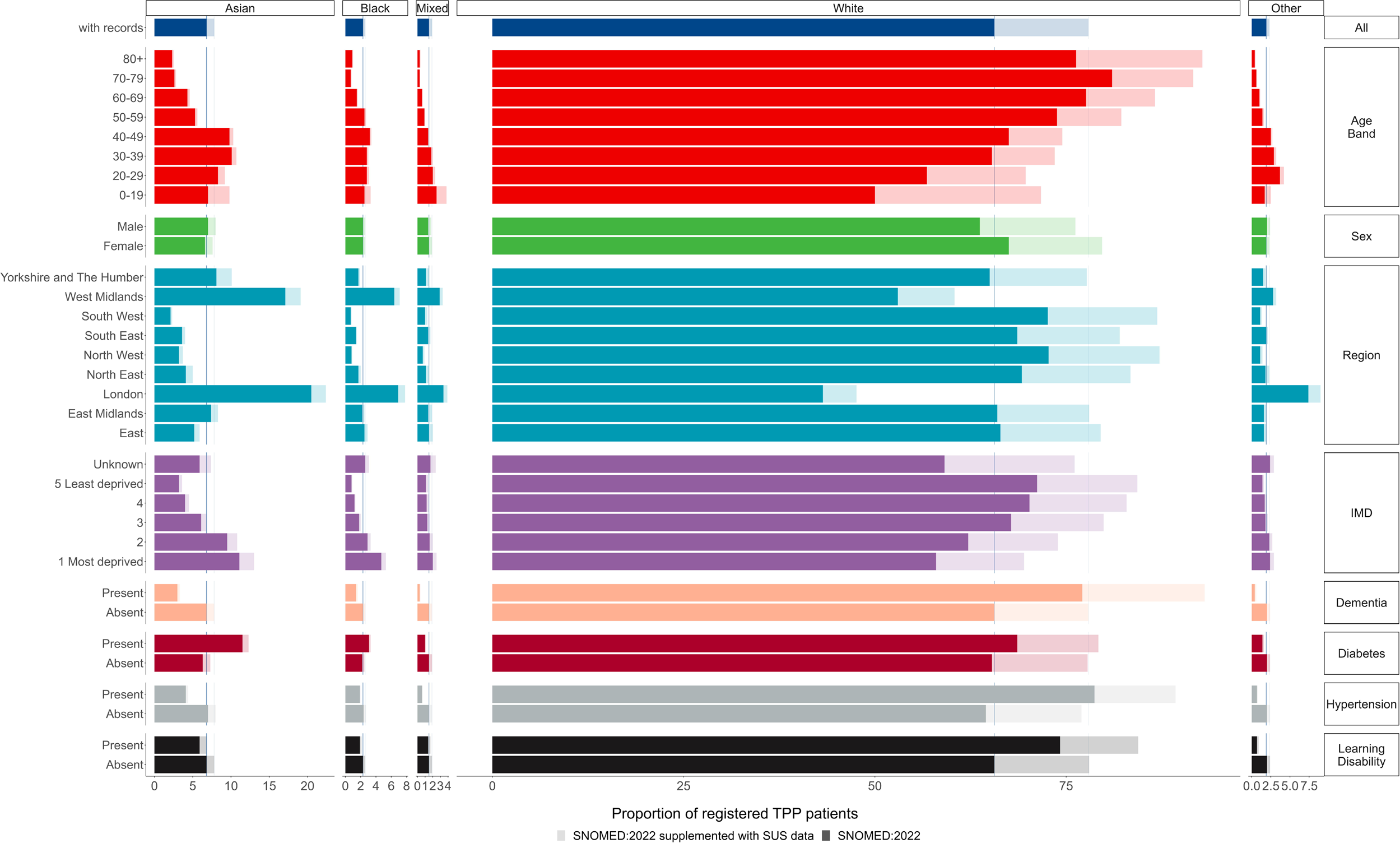

### Consistency of ethnicity recording within patients

3.1% (611,260) of the 19,618,135 patients with a recorded ethnicity had at least one ethnicity record that was discordant with the latest recorded ethnicity (Table 3). Patients whose latest recorded ethnicity was categorised as Mixed were most likely to have a discordant ethnicity recording (32.2%, 118,560); of whom 17.0% (62,565) also had a recorded ethnicity of White. 5.7% (33,205) of the 583,770 patients with the latest recorded ethnicity of Black also had a recorded ethnicity of White (Table 1).

**Table 1:** Count of patients with at least one recording of each ethnicity (proportion of latest ethnicity).

| Latest ethnicity | Any recorded ethnicity |  |  |  |  | Any discordant ethnicity |
| --- | --- | --- | --- | --- | --- | --- |
|  | Asian | Black | Mixed | White | Other |  |
| Asian:<br>1,708,430 | 1,708,430<br>(100.0) | 8,640<br>(0.5) | 25,955<br>(1.5) | 42,760<br>(2.5) | 41,175<br>(2.4) | 109,060<br>(6.4) |
| Black:<br>583,770 | 6,680<br>(1.1) | 583,770<br>(100.0) | 41,245<br>(7.1) | 33,205<br>(5.7) | 11,495<br>(2.0) | 85,075<br>(14.6) |
| Mixed:<br>367,980 | 18,400<br>(5.0) | 32,990<br>(9.0) | 367,980<br>(100.0) | 62,565<br>(17.0) | 15,920<br>(4.3) | 118,560<br>(32.2) |
| White:<br>16,468,610 | 31,635<br>(0.2) | 25,115<br>(0.2) | 62,030<br>(0.4) | 16,468,610<br>(100.0) | 81,875<br>(0.5) | 189,020<br>(1.1) |
| Other:<br>489,350 | 32,875<br>(6.7) | 9,430<br>(1.9) | 16,795<br>(3.4) | 60,865<br>(12.4) | 489,350<br>(100.0) | 109,545<br>(22.4) |

Overall, for 19,364,120 (98.7%) of patients, their latest recorded ethnicity in primary care matched their most frequently recorded ethnicity in primary care (Table 2). 16,390,425 (99.5%) patients with the most recent ethnicity ‘White’ had matching most frequently recorded ethnicity. Other was the least concordant group, just 81.6% (399,440) of patients with the most recent ethnicity ‘Mixed’ had matching most frequently recorded ethnicity. 0.9% (5,450) of patients with latest ethnicity ‘Black’ had the most frequently recorded ethnicity ‘White’ (Figure 3, Extended Data Table 4).

**Table 2:**
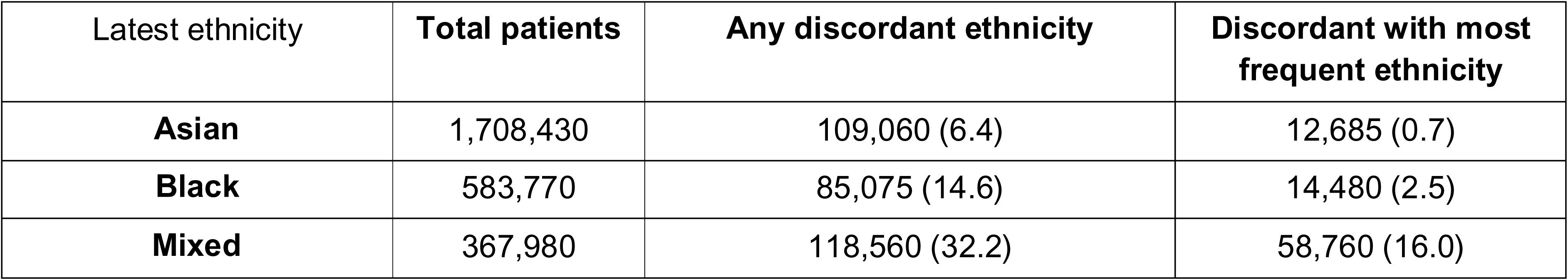
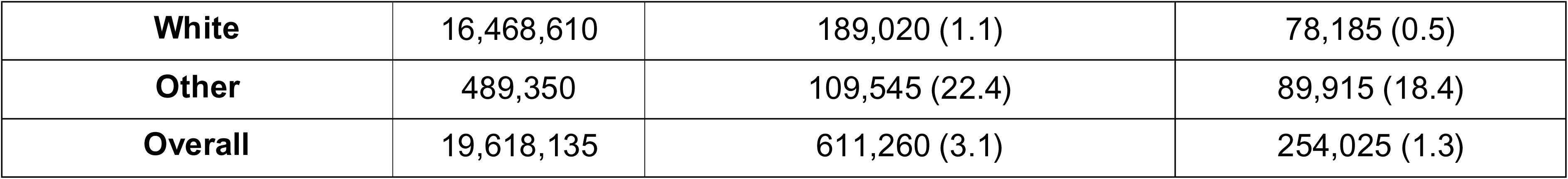
Count of patients with any recorded discordant ethnicity and a discordant ‘most frequently recorded’ ethnicity in primary care, according to latest ethnicity.

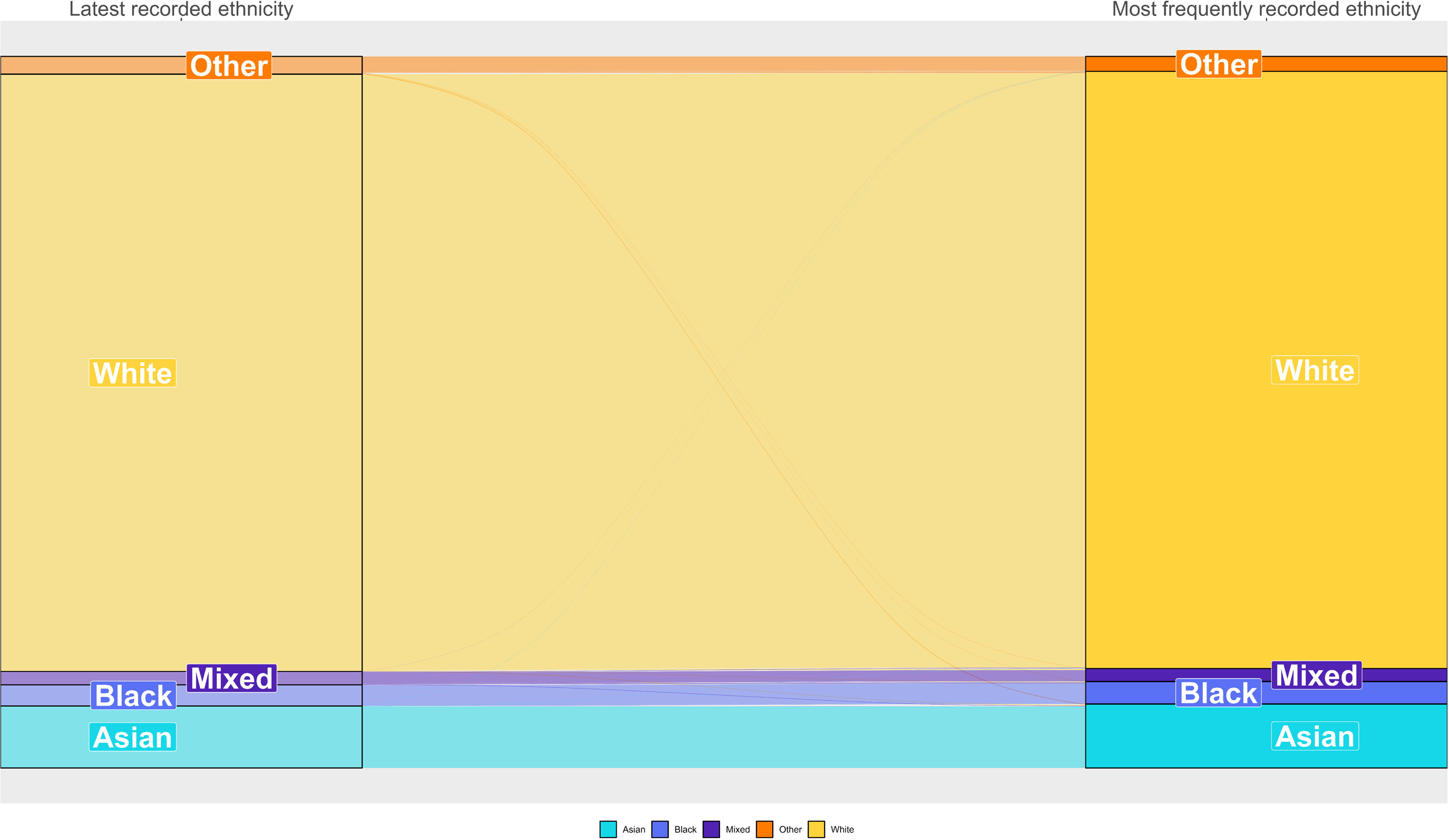

### Consistency of ethnicity recording across data sources (primary care versus secondary care)

Of the 19.6 million total patients with a primary care ethnicity record, 12.9 million (66.0%) also had a secondary care ethnicity record. The proportion of patients with no secondary care coded ethnicity ranged from 31.9% in the White group to 58.6% in the Other group (Extended Data Table 5). SNOMED:2022 and secondary care coded ethnicity matched for 93.5% of patients with both coded ethnicities, ranging from 34.8% in the Mixed group to 96.9% in the White group (Figure 4, Extended Data Table 6).

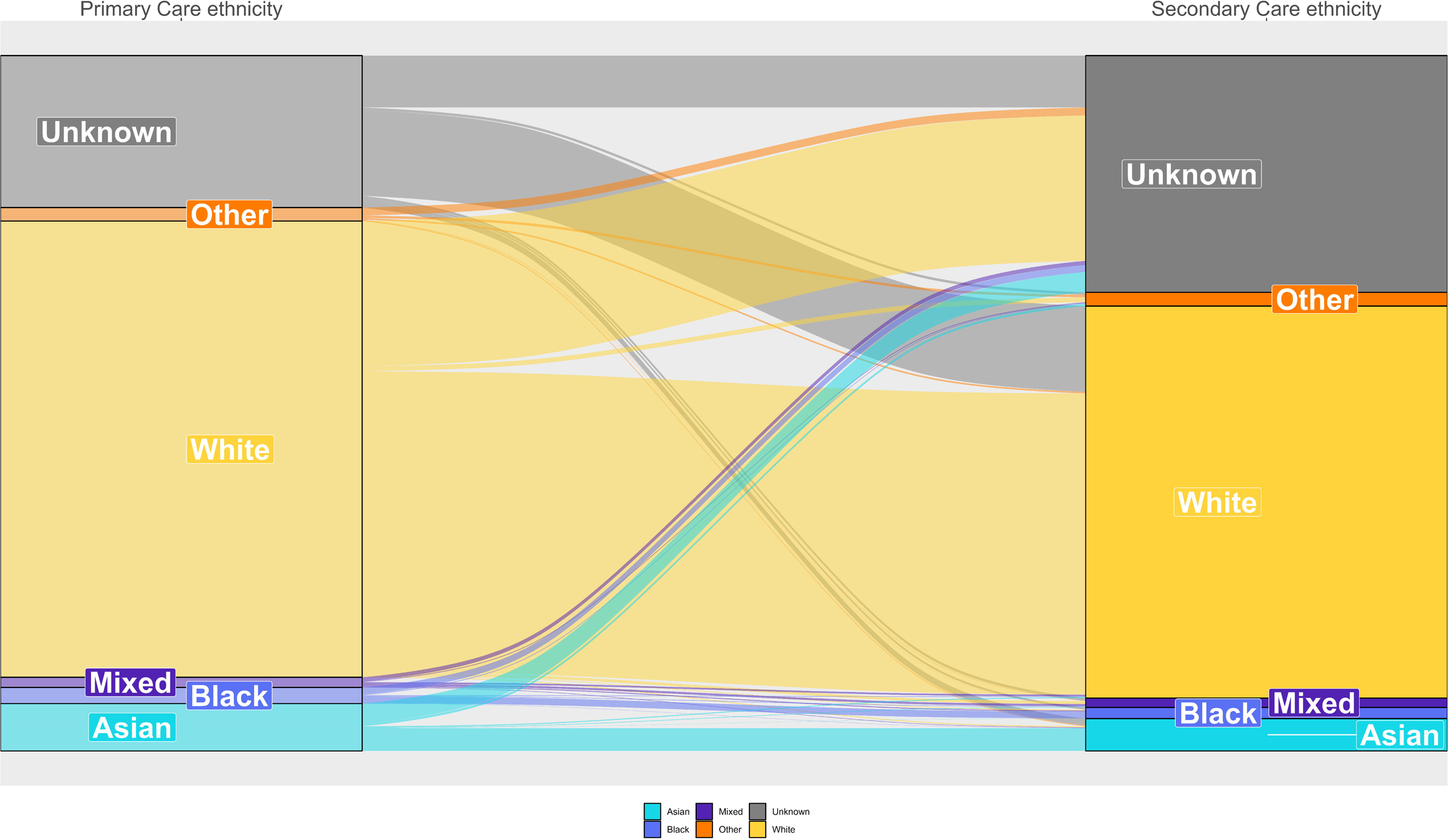

### Comparison with the 2021 UK census population

The proportion of patients in each ethnicity group based on primary care records as of January 2022 was within 2.9 percentage points of the 2021 Census estimate for the same ethnicity group across England as a whole (Asian: 8.7% primary care, 8.8% Census; Black: 3.0%, 4.2%; Mixed: 1.9%, 3.0%; White: 84.0%, 81.0%; Other: 2.5%, 2.9%). When supplemented secondary care data this increased to 3.2% (Figure 5, Extended Data Table 7). In primary care records the White population was underrepresented in all regions other than the North West (7.1% percentage points higher than Census estimates), South East (2.8%) and South West (0.6%) and was most severely underestimated in the West Midlands (−12.5%). The Asian population was overrepresented in all regions other than the North West (−3.6%) and South East (−1.6%) (Figure 6, Extended Data Table 7). We also compared the primary care data to the 2021 Census estimates using 2021 rather than 2001 ethnicity groups (Extended Data Figures 1,2 and Extended Data Table 8).

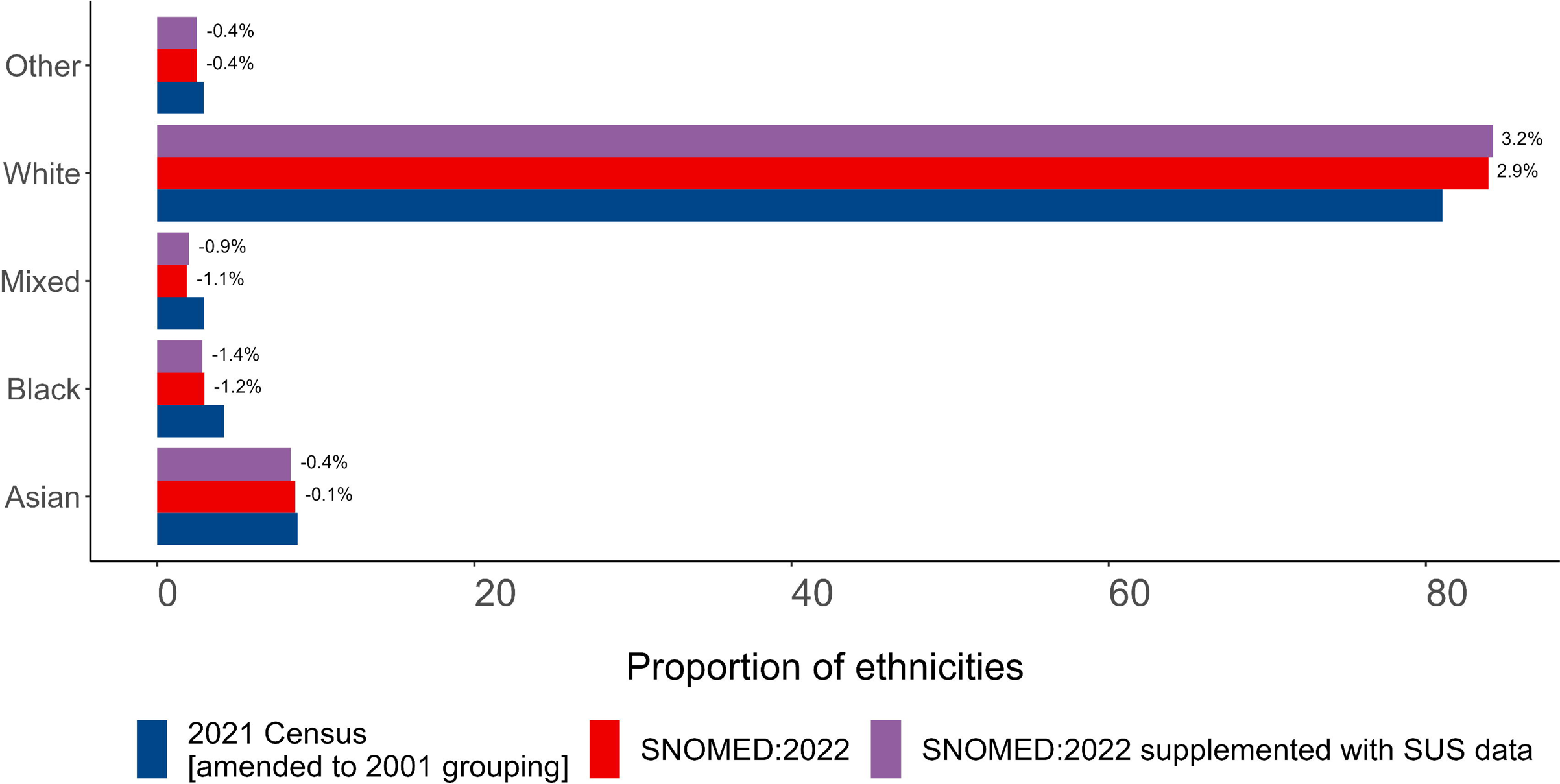

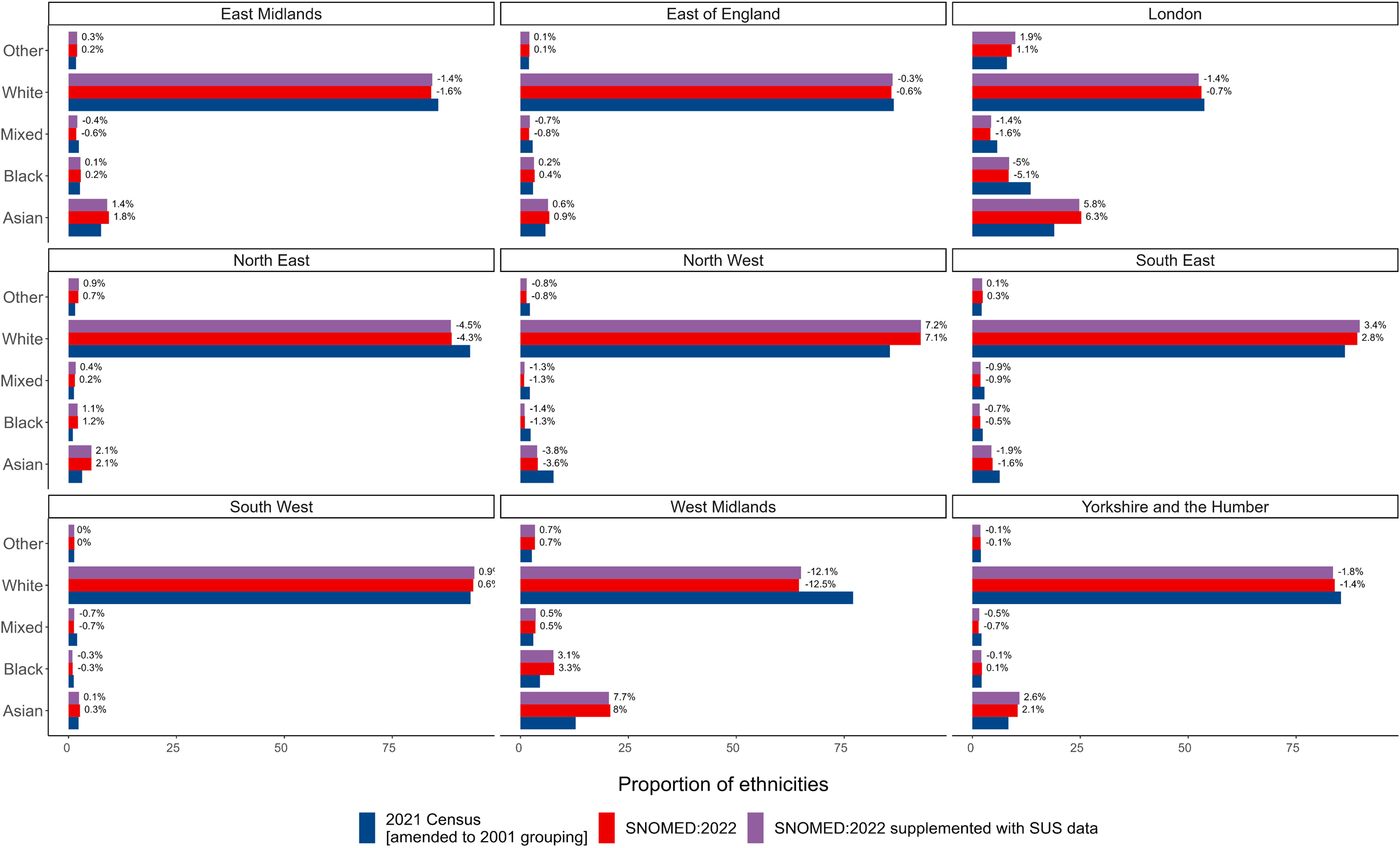

## Discussion

### Summary

This study reported ethnicity recording quality in around 25 million patients registered with a general practice in England and available for analysis in the OpenSAFELY-TPP database. Over three quarters of all patients had at least one ethnicity record in primary care data. When supplemented with hospital records, ethnicity recording was 92.5% complete, which is consistent with previously reported England-wide primary care data sources^28,29^. The reported concordance of primary and secondary care records of 93.5% is consistent with those previously reported^30^. Despite regional variations the overall ethnicity breakdown across all English OpenSAFELY-TPP practices was similar to the 2021 Census.

### Strengths and weaknesses

This study provides a breakdown of primary care coding in OpenSAFELY-TPP by key clinical and demographic characteristics. The key strengths of this study are the use of large EHR datasets representing roughly 40% of the population of England registered with a GP, which enabled us to assess the quality of ethnicity data against a variety of important clinical characteristics.

Practices may utilise differing strategies for collecting ethnicity information from patients. Typically ethnicity is self-reported by the patient at registration or during consultation^31^ but may not always be self-reported. OpenSAFELY-TPP was missing ethnicity for 21.8% of patients, and the missingness of ethnicity data in EHRs may not be random^6^.

This study focussed on the 5 Group ethnicity of the SNOMED:2022 codelists categorisation. However, there can be important variations in clinical care within these broad categories, as seen in COVID vaccine uptake^32,33^.

It is common for OpenSAFELY-TPP studies to supplement the primary care recorded ethnicity, where missing, with ethnicity data from secondary care^10,11,34^. The representativeness of the CTV3:2020 coded ethnicity supplemented with SUS data has been reported previously^34^. However, secondary care data is only available for people attending hospital within the time period that data were available (currently April 2019 onwards in OpenSAFELY). The population who still have no ethnicity record after supplementation are likely very different to the wider population, for example having a much lower chance of having been admitted to hospital, or interacting with healthcare services generally.

This study represents a snapshot of ethnicity recording as of 1 January 2022, and does not provide insights into temporal trends in ethnicity recording. Trends in ethnicity recording over time are difficult to investigate due to loss of record date during transfer of clinical records when patients register with a new practice (Extended Data Figure 3). Therefore, we are unable to assess the impact of QOF financial incentives being rescinded in 2011/12.

The most up-to-date formal estimates of England’s population by ethnic group currently available are from the 2021 Census. Accuracy of the 2021 Census ethnicity estimates may vary by region. The 2021 census response rate was not even between regions, ranging from 95% in London to 98% in the South East, South West and East of England^35^. The 2021 census used multiple imputation to account for missing ethnicity^36^, the percentage of eligible persons who had an ethnicity value imputed or edited was not even between regions. Imputation rate was highest in London (2.0%) and lowest in the North East (1.0%)^35^.

There are limitations in comparing the GP-registered population with the census population as differences naturally arise. For example, patients registered with a GP may have left the country some years ago and hence not be counted in the census; certain populations are less likely to be registered with a GP (such as Gypsy, Roma and Traveller communities^37^ and migrants^38,39^); not everyone responds to the census but some may be registered with a GP; and regional differences occur, for example due to students moving to cities during term-time. We looked at the GP-registered population in January 2022 whereas the census was taken in March 2021 therefore some small changes in population also may have occurred during this time.

### Findings in Context

Over 20 studies have been conducted using the OpenSAFELY framework. It is important to understand the data issues with using ethnicity in OpenSAFELY. Whilst ethnicity data has been shown to be more complete for the CTV3:2020 codelist than the SNOMED:2022 codelist^13^, the CTV3:2020 codelist included codes such as “XaJSe: Muslim - ethnic category 2001 census” which relate to religion rather than ethnicity and were, therefore, excluded from the SNOMED:2022 codelist. The common practice of supplementing CTV3:2020 coded ethnicity with either secondary care data or the PRIMIS codelists could lead to inconsistent classification as both secondary care data and PRIMIS codelists follow the 2001 census categories.

Recording ethnicity is not straightforward. Indeed, despite often being used as a key variable to describe health, the idea of “ethnicity” has been disputed^40^. Ethnicity is a complex mixture of social constructs, genetic make-up, and cultural identity ^41^. Self-identified ethnicity is not a fixed concept and evolving socio-cultural trends could contribute to changes in a person’s self-identified ethnic group, particularly for those with mixed heritage^42^. It is therefore perhaps not surprising to see lower levels of concordance between latest ethnicity and most common ethnicity in those with latest ethnicity coded as ‘mixed’. It is not clear to what extent this would explain all the discordance we identified or whether other factors such as data error are involved. Our findings agree with previous literature, both from the US and UK^5,42^, which suggest that the consistency of ethnicity information tends to be highest for white populations, and lowest for Mixed or Other racial/ethnic groups^43^.

The 2001 census categories are the NHS standard for ethnicity^17^, but we have not been able to find any explanation for the continued use of the 2001 census categories as the standard.

Due to the significant differences experienced by ethnic groups in terms of health outcomes, accurate ethnicity coding to the most granular code possible is crucial. Although we have focussed on codelist categorisations based on the 2001 census categories, ethnicity can be extracted for each of the component codes (Extended Data Table 8) so researchers have the option to use custom categorisations as required.

We believe that the SNOMED:2022 codelist and codelist categorisation provides a more consistent representation of ethnicity as defined by the 2001 census categories than the CTV3:2020 codelist, and should be the preferred codelist and categorisation for primary care ethnicity.

### Policy Implications and Interpretation

This paper is principally to inform interpretation of the numerous current and future analyses completed and published using OpenSAFELY-TPP and similar UK electronic healthcare databases. The practice of supplementing primary care ethnicity with secondary care ethnicity from SUS can, depending on the study design, introduce bias and should be used with caution. For example, patients who have more clinical interactions are more likely to have a recorded ethnicity and therefore patients with a recorded ethnicity in secondary care data may tend to be sicker than the general population. Ethnicity recording has been found to be more complete for patients who died in hospital compared with those discharged^5^.

### Conclusions

This report describes the completeness and consistency of primary care ethnicity in OpenSAFELY-TPP and suggests the adoption of the SNOMED:2022 codelist and codelist categorisation as the best standard method.

## Data Availability

Access to the underlying identifiable and potentially re-identifiable pseudonymised electronic health record data is tightly governed by various legislative and regulatory frameworks, and restricted by best practice. The data in OpenSAFELY is drawn from General Practice data across England where TPP is the Data Processor. TPP developers (CB, JC, JP, FH, and SH) initiate an automated process to create pseudonymised records in the core OpenSAFELY database, which are copies of key structured data tables in the identifiable records. These are linked onto key external data resources that have also been pseudonymised via SHA-512 one-way hashing of NHS numbers using a shared salt. Bennett Institute for Applied Data Science developers and PIs (BG, CEM, SB, AJW, KW, WJH, HJC, DE, PI, SD, GH, BBC, RMS, ID, TW, TO, SM, CLS, LB, JM, MW, RYP, KB, EJW and CTR) holding contracts with NHS England have access to the OpenSAFELY pseudonymised data tables as needed to develop the OpenSAFELY tools. These tools in turn enable researchers with OpenSAFELY Data Access Agreements to write and execute code for data management and data analysis without direct access to the underlying raw pseudonymised patient data, and to review the outputs of this code. All code for the full data management pipeline-from raw data to completed results for this analysis-and for the OpenSAFELY platform as a whole is available for review at github.com/OpenSAFELY.

## Administrative

## Acknowledgements

We are very grateful for all the support received from the TPP Technical Operations team throughout this work, and for generous assistance from the information governance and database teams at NHS England / NHSX.

## Information governance and ethical approval

NHS England is the data controller; TPP is the data processor; and the researchers on OpenSAFELY are acting with the approval of NHS England. This implementation of OpenSAFELY is hosted within the TPP environment which is accredited to the ISO 27001 information security standard and is NHS IG Toolkit compliant;^44,45^ patient data has been pseudonymised for analysis and linkage using industry standard cryptographic hashing techniques; all pseudonymised datasets transmitted for linkage onto OpenSAFELY are encrypted; access to the platform is via a virtual private network (VPN) connection, restricted to a small group of researchers; the researchers hold contracts with NHS England and only access the platform to initiate database queries and statistical models; all database activity is logged; only aggregate statistical outputs leave the platform environment following best practice for anonymisation of results such as statistical disclosure control for low cell counts.^46^ The OpenSAFELY research platform adheres to the obligations of the UK General Data Protection Regulation (GDPR) and the Data Protection Act 2018. In March 2020, the Secretary of State for Health and Social Care used powers under the UK Health Service (Control of Patient Information) Regulations 2002 (COPI) to require organisations to process confidential patient information for the purposes of protecting public health, providing healthcare services to the public and monitoring and managing the COVID-19 outbreak and incidents of exposure; this sets aside the requirement for patient consent.^47^ Taken together, these provide the legal bases to link patient datasets on the OpenSAFELY platform. GP practices, from which the primary care data are obtained, are required to share relevant health information to support the public health response to the pandemic, and have been informed of the OpenSAFELY analytics platform.

This study was approved by the Health Research Authority (REC reference 20/LO/0651) and by the LSHTM Ethics Board (reference 21863).

## Data access and verification

Access to the underlying identifiable and potentially re-identifiable pseudonymised electronic health record data is tightly governed by various legislative and regulatory frameworks, and restricted by best practice. The data in OpenSAFELY is drawn from General Practice data across England where TPP is the Data Processor. TPP developers (CB, JC, JP, FH, and SH) initiate an automated process to create pseudonymised records in the core OpenSAFELY database, which are copies of key structured data tables in the identifiable records. These are linked onto key external data resources that have also been pseudonymised via SHA-512 one-way hashing of NHS numbers using a shared salt. Bennett Institute for Applied Data Science developers and PIs (BG, CEM, SB, AJW, KW, WJH, HJC, DE, PI, SD, GH, BBC, RMS, ID, KB, EJW and CTR) holding contracts with NHS England have access to the OpenSAFELY pseudonymised data tables as needed to develop the OpenSAFELY tools. These tools in turn enable researchers with OpenSAFELY Data Access Agreements to write and execute code for data management and data analysis without direct access to the underlying raw pseudonymised patient data, and to review the outputs of this code. All code for the full data management pipeline—from raw data to completed results for this analysis—and for the OpenSAFELY platform as a whole is available for review at github.com/OpenSAFELY.

The data management and analysis code for this paper was led by CA and RP and contributed to by WJH, LH, and AJW.

Guarantor

BG is guarantor.

