## Extended Data Table 1 for "Consistency, completeness and external validity of ethnicity recording in NHS primary care records: a cohort study in 25 million patients’ records at source using OpenSAFELY"

Table 1: Count of patients with a recorded ethnicity in OpenSAFELY-TPP (proportion of registered TPP population) by clinical and demographic subgroups. All counts are rounded to the nearest 5.

| | SNOMED 2022 | SNOMED 2022 with SUS data | Population | Percentage point increase with SUS data |
|---|---|---|---|---|
| all | | | | |
| with records | 19,618,135 (78.2) | 23,228,760 (92.5) | 25,102,210 | (14.3) |
| age band | | | | |
| 0-19 | 3,506,695 (63.6) | 5,019,200 (91.1) | 5,509,975 | (27.5) |
| 20-29 | 2,300,970 (73.6) | 2,767,405 (88.5) | 3,128,250 | (14.9) |
| 30-39 | 2,985,455 (83.0) | 3,327,535 (92.5) | 3,597,050 | (9.5) |
| 40-49 | 2,730,080 (84.5) | 2,989,030 (92.6) | 3,228,995 | (8.1) |
| 50-59 | 2,887,720 (83.9) | 3,199,355 (93.0) | 3,440,755 | (9.1) |
| 60-69 | 2,326,765 (84.9) | 2,589,095 (94.5) | 2,741,010 | (9.6) |
| 70-79 | 1,875,495 (85.2) | 2,118,495 (96.2) | 2,201,400 | (11.0) |
| 80+ | 1,004,955 (80.1) | 1,218,635 (97.1) | 1,254,775 | (17.0) |
| sex | | | | |
| Female | 10,005,930 (79.8) | 11,782,895 (94.0) | 12,532,940 | (14.2) |
| Male | 9,612,205 (76.5) | 11,445,860 (91.1) | 12,569,270 | (14.6) |
| region | | | | |
| East | 4,488,910 (77.3) | 5,355,465 (92.2) | 5,808,670 | (14.9) |
| East Midlands | 3,413,070 (78.6) | 4,020,145 (92.6) | 4,343,010 | (14.0) |
| London | 1,460,750 (81.4) | 1,629,015 (90.7) | 1,795,300 | (9.3) |
| North East | 914,095 (78.0) | 1,103,540 (94.2) | 1,171,695 | (16.2) |
| North West | 1,693,835 (78.4) | 2,029,735 (94.0) | 2,159,325 | (15.6) |
| South East | 1,265,915 (77.0) | 1,502,860 (91.4) | 1,644,505 | (14.4) |
| South West | 2,697,960 (77.4) | 3,219,590 (92.4) | 3,483,925 | (15.0) |
| West Midlands | 837,230 (82.2) | 947,725 (93.0) | 1,018,680 | (10.8) |
| Yorkshire and The Humber | 2,816,935 (77.4) | 3,385,170 (93.0) | 3,638,855 | (15.6) |
| IMD | | | | |
| 1 Most deprived | 3,885,360 (78.2) | 4,633,110 (93.3) | 4,968,330 | (15.1) |
| 2 | 3,821,805 (78.5) | 4,515,210 (92.8) | 4,868,115 | (14.3) |
| 3 | 4,027,315 (78.9) | 4,724,895 (92.6) | 5,102,160 | (13.7) |
| 4 | 3,871,300 (78.3) | 4,559,785 (92.3) | 4,942,265 | (14.0) |
| 5 Least deprived | 3,482,535 (77.7) | 4,116,455 (91.8) | 4,481,755 | (14.1) |
| Unknown | 529,815 (71.6) | 679,300 (91.8) | 739,585 | (20.2) |
| dementia | | | | |
| Present | 36,645 (82.2) | 44,075 (98.9) | 44,560 | (16.7) |
| Absent | 19,581,490 (78.1) | 23,184,680 (92.5) | 25,057,655 | (14.4) |
| diabetes | | | | |
| Present | 2,049,495 (85.6) | 2,335,080 (97.5) | 2,394,370 | (11.9) |
| Absent | 17,568,640 (77.4) | 20,893,675 (92.0) | 22,707,845 | (14.6) |
| hypertension | | | | |
| Present | 1,621,375 (86.0) | 1,833,885 (97.2) | 1,886,165 | (11.2) |
| Absent | 17,996,755 (77.5) | 21,394,870 (92.2) | 23,216,045 | (14.7) |
| learning disability | | | | |
| Present | 120,385 (84.2) | 137,380 (96.1) | 143,025 | (11.9) |
| Absent | 19,497,750 (78.1) | 23,091,375 (92.5) | 24,959,185 | (14.4) |
