## Extended Data Table 2 for "Consistency, completeness and external validity of ethnicity recording in NHS primary care records: a cohort study in 25 million patients’ records at source using OpenSAFELY"

Table 2: Count of patients with a recorded ethnicity in OpenSAFELY TPP by ethnicity group (proportion of registered TPP population) and clinical and demographic subgroups. All counts are rounded to the nearest 5.

| | | Asian | | Black | | Mixed | | White | | Other | |
|---|---|---|---|---|---|---|---|---|---|---|---|
| | | **SNOMED 2022** | **SNOMED 2022 with SUS data** | **SNOMED 2022** | **SNOMED 2022 with SUS data** | **SNOMED 2022** | **SNOMED 2022 with SUS data** | **SNOMED 2022** | **SNOMED 2022 with SUS data** | **SNOMED 2022** | **SNOMED 2022 with SUS data** |
| all | with records | 1,708,430 (6.8) | 1,955,095 (7.8) | 583,770 (2.3) | 659,410 (2.6) | 367,980 (1.5) | 466,220 (1.9) | 16,468,610 (65.6) | 19,566,635 (77.9) | 489,350 (1.9) | 581,395 (2.3) |
| age band | 0-19 | 383,285 (7.0) | 536,695 (9.7) | 138,355 (2.5) | 182,015 (3.3) | 137,175 (2.5) | 210,360 (3.8) | 2,754,320 (50.0) | 3,951,305 (71.7) | 93,560 (1.7) | 138,830 (2.5) |
| | 20-29 | 259,930 (8.3) | 289,485 (9.3) | 86,955 (2.8) | 95,870 (3.1) | 63,215 (2.0) | 72,340 (2.3) | 1,775,885 (56.8) | 2,180,550 (69.7) | 114,985 (3.7) | 129,165 (4.1) |
| | 30-39 | 363,460 (10.1) | 386,210 (10.7) | 102,485 (2.8) | 109,505 (3.0) | 64,395 (1.8) | 70,745 (2.0) | 2,350,510 (65.3) | 2,644,670 (73.5) | 104,605 (2.9) | 116,405 (3.2) |
| | 40-49 | 315,825 (9.8) | 331,920 (10.3) | 104,610 (3.2) | 109,780 (3.4) | 46,475 (1.4) | 49,975 (1.5) | 2,181,060 (67.5) | 2,407,950 (74.6) | 82,105 (2.5) | 89,405 (2.8) |
| | 50-59 | 182,030 (5.3) | 192,220 (5.6) | 85,405 (2.5) | 90,855 (2.6) | 31,895 (0.9) | 34,685 (1.0) | 2,539,560 (73.8) | 2,827,325 (82.2) | 48,835 (1.4) | 54,280 (1.6) |
| | 60-69 | 116,945 (4.3) | 124,535 (4.5) | 40,280 (1.5) | 43,280 (1.6) | 15,365 (0.6) | 16,995 (0.6) | 2,126,970 (77.6) | 2,373,055 (86.6) | 27,200 (1.0) | 31,230 (1.1) |
| | 70-79 | 58,290 (2.6) | 62,645 (2.8) | 14,745 (0.7) | 15,960 (0.7) | 6,290 (0.3) | 7,285 (0.3) | 1,783,385 (81.0) | 2,017,300 (91.6) | 12,785 (0.6) | 15,305 (0.7) |
| | 80+ | 28,670 (2.3) | 31,390 (2.5) | 10,930 (0.9) | 12,145 (1.0) | 3,165 (0.3) | 3,840 (0.3) | 956,915 (76.3) | 1,164,480 (92.8) | 5,270 (0.4) | 6,780 (0.5) |
| sex | Female | 827,980 (6.6) | 949,050 (7.6) | 291,010 (2.3) | 328,460 (2.6) | 186,680 (1.5) | 234,280 (1.9) | 8,461,075 (67.5) | 9,987,690 (79.7) | 239,185 (1.9) | 283,420 (2.3) |
| | Male | 880,450 (7.0) | 1,006,045 (8.0) | 292,760 (2.3) | 330,955 (2.6) | 181,300 (1.4) | 231,940 (1.8) | 8,007,535 (63.7) | 9,578,950 (76.2) | 250,160 (2.0) | 297,980 (2.4) |
| region | East | 300,605 (5.2) | 342,735 (5.9) | 147,865 (2.5) | 168,770 (2.9) | 89,825 (1.5) | 116,180 (2.0) | 3,856,555 (66.4) | 4,617,355 (79.5) | 94,055 (1.6) | 110,425 (1.9) |
| | East Midlands | 319,445 (7.4) | 360,305 (8.3) | 97,320 (2.2) | 110,695 (2.5) | 60,985 (1.4) | 81,860 (1.9) | 2,867,445 (66.0) | 3,386,935 (78.0) | 67,880 (1.6) | 80,350 (1.9) |
| | London | 368,555 (20.5) | 403,495 (22.5) | 123,080 (6.9) | 138,875 (7.7) | 60,995 (3.4) | 70,810 (3.9) | 775,090 (43.2) | 853,735 (47.6) | 133,035 (7.4) | 162,100 (9.0) |
| | North East | 48,500 (4.1) | 58,495 (5.0) | 19,950 (1.7) | 22,990 (2.0) | 13,465 (1.1) | 18,160 (1.5) | 811,235 (69.2) | 977,415 (83.4) | 20,940 (1.8) | 26,485 (2.3) |
| | North West | 68,150 (3.2) | 78,725 (3.6) | 17,470 (0.8) | 19,705 (0.9) | 14,420 (0.7) | 19,055 (0.9) | 1,570,265 (72.7) | 1,882,895 (87.2) | 23,535 (1.1) | 29,350 (1.4) |
| | South East | 59,670 (3.6) | 66,180 (4.0) | 23,510 (1.4) | 25,550 (1.6) | 23,715 (1.4) | 28,355 (1.7) | 1,128,545 (68.6) | 1,348,710 (82.0) | 30,470 (1.9) | 34,070 (2.1) |
| | South West | 71,795 (2.1) | 78,600 (2.3) | 25,735 (0.7) | 28,730 (0.8) | 33,950 (1.0) | 42,945 (1.2) | 2,529,835 (72.6) | 3,026,520 (86.9) | 36,640 (1.1) | 42,795 (1.2) |
| | West Midlands | 174,370 (17.1) | 194,405 (19.1) | 65,400 (6.4) | 72,195 (7.1) | 29,220 (2.9) | 33,420 (3.3) | 540,185 (53.0) | 615,885 (60.5) | 28,055 (2.8) | 31,815 (3.1) |
| | Yorkshire and The Humber | 294,930 (8.1) | 369,235 (10.1) | 62,195 (1.7) | 70,370 (1.9) | 40,435 (1.1) | 54,080 (1.5) | 2,365,245 (65.0) | 2,828,245 (77.7) | 54,135 (1.5) | 63,245 (1.7) |
| IMD | 1 Most deprived | 552,360 (11.1) | 646,705 (13.0) | 232,370 (4.7) | 262,400 (5.3) | 100,090 (2.0) | 127,030 (2.6) | 2,882,250 (58.0) | 3,452,775 (69.5) | 118,290 (2.4) | 144,200 (2.9) |
| | 2 | 460,265 (9.5) | 521,460 (10.7) | 143,240 (2.9) | 161,860 (3.3) | 79,195 (1.6) | 99,685 (2.0) | 3,029,195 (62.2) | 3,600,725 (74.0) | 109,910 (2.3) | 131,485 (2.7) |
| | 3 | 312,525 (6.1) | 349,530 (6.9) | 91,495 (1.8) | 102,700 (2.0) | 68,860 (1.3) | 86,075 (1.7) | 3,460,745 (67.8) | 4,076,195 (79.9) | 93,695 (1.8) | 110,400 (2.2) |
| | 4 | 197,745 (4.0) | 222,365 (4.5) | 59,885 (1.2) | 67,155 (1.4) | 59,330 (1.2) | 74,685 (1.5) | 3,468,920 (70.2) | 4,096,445 (82.9) | 85,420 (1.7) | 99,135 (2.0) |
| | 5 Least deprived | 142,175 (3.2) | 160,720 (3.6) | 37,805 (0.8) | 42,430 (0.9) | 48,195 (1.1) | 61,140 (1.4) | 3,190,305 (71.2) | 3,777,735 (84.3) | 64,055 (1.4) | 74,435 (1.7) |
| | Unknown | 43,360 (5.9) | 54,320 (7.3) | 18,970 (2.6) | 22,865 (3.1) | 12,310 (1.7) | 17,610 (2.4) | 437,195 (59.1) | 562,760 (76.1) | 17,980 (2.4) | 21,740 (2.9) |
| dementia | Present | 1,325 (3.0) | 1,455 (3.3) | 625 (1.4) | 705 (1.6) | 155 (0.3) | 180 (0.4) | 34,360 (77.1) | 41,485 (93.1) | 180 (0.4) | 250 (0.6) |
| | Absent | 1,707,105 (6.8) | 1,953,640 (7.8) | 583,140 (2.3) | 658,705 (2.6) | 367,820 (1.5) | 466,040 (1.9) | 16,434,250 (65.6) | 19,525,150 (77.9) | 489,170 (2.0) | 581,145 (2.3) |
| diabetes | Present | 275,225 (11.5) | 294,210 (12.3) | 73,500 (3.1) | 78,875 (3.3) | 23,905 (1.0) | 26,450 (1.1) | 1,643,120 (68.6) | 1,896,130 (79.2) | 33,740 (1.4) | 39,415 (1.6) |
| | Absent | 1,433,205 (6.3) | 1,660,885 (7.3) | 510,265 (2.2) | 580,540 (2.6) | 344,075 (1.5) | 439,770 (1.9) | 14,825,490 (65.3) | 17,670,505 (77.8) | 455,605 (2.0) | 541,980 (2.4) |
| hypertension | Present | 77,420 (4.1) | 82,950 (4.4) | 35,695 (1.9) | 38,290 (2.0) | 11,145 (0.6) | 12,420 (0.7) | 1,483,585 (78.7) | 1,684,145 (89.3) | 13,535 (0.7) | 16,085 (0.9) |
| | Absent | 1,631,010 (7.0) | 1,872,140 (8.1) | 548,075 (2.4) | 621,125 (2.7) | 356,835 (1.5) | 453,800 (2.0) | 14,985,025 (64.5) | 17,882,495 (77.0) | 475,815 (2.0) | 565,315 (2.4) |
| learning disability | Present | 8,450 (5.9) | 9,750 (6.8) | 2,745 (1.9) | 3,085 (2.2) | 2,000 (1.4) | 2,450 (1.7) | 106,145 (74.2) | 120,710 (84.4) | 1,040 (0.7) | 1,385 (1.0) |
| | Absent | 1,699,980 (6.8) | 1,945,345 (7.8) | 581,020 (2.3) | 656,325 (2.6) | 365,980 (1.5) | 463,770 (1.9) | 16,362,460 (65.6) | 19,445,925 (77.9) | 488,310 (2.0) | 580,010 (2.3) |
