## Extended Data Table 3 for "Consistency, completeness and external validity of ethnicity recording in NHS primary care records: a cohort study in 25 million patients’ records at source using OpenSAFELY"

Table 3: Count of patients with a recorded ethnicity in OpenSAFELY TPP by ethnicity group (proportion of registered TPP population) and clinical and demographic subgroups. All counts are rounded to the nearest 5.

| | | Indian | | Pakistani | | Bangladeshi | | Other Asian | | Caribbean | | African | | Other Black | | White and Black Caribbean | |
|---|---|---|---|---|---|---|---|---|---|---|---|---|---|---|---|---|---|
| | | **SNOMED 2022** | **SNOMED 2022 with SUS data** | **SNOMED 2022** | **SNOMED 2022 with SUS data** | **SNOMED 2022** | **SNOMED 2022 with SUS data** | **SNOMED 2022** | **SNOMED 2022 with SUS data** | **SNOMED 2022** | **SNOMED 2022 with SUS data** | **SNOMED 2022** | **SNOMED 2022 with SUS data** | **SNOMED 2022** | **SNOMED 2022 with SUS data** | **SNOMED 2022** | **SNOMED 2022 with SUS data** |
| all | with records | 690,425 (2.8) | 765,870 (3.1) | 499,570 (2.0) | 604,255 (2.4) | 120,610 (0.5) | 139,920 (0.6) | 397,825 (1.6) | 445,050 (1.8) | 113,575 (0.5) | 132,025 (0.5) | 369,315 (1.5) | 410,730 (1.6) | 100,880 (0.4) | 116,705 (0.5) | 82,660 (0.3) | 105,695 (0.4) |
| age band | 0-19 | 123,800 (2.2) | 169,220 (3.1) | 143,275 (2.6) | 211,650 (3.8) | 34,080 (0.6) | 46,840 (0.9) | 82,130 (1.5) | 108,970 (2.0) | 15,525 (0.3) | 22,640 (0.4) | 95,135 (1.7) | 124,240 (2.3) | 27,690 (0.5) | 35,135 (0.6) | 30,755 (0.6) | 47,570 (0.9) |
| | 20-29 | 101,930 (3.3) | 108,965 (3.5) | 74,360 (2.4) | 88,700 (2.8) | 19,345 (0.6) | 21,455 (0.7) | 64,300 (2.1) | 70,410 (2.3) | 12,965 (0.4) | 15,680 (0.5) | 58,895 (1.9) | 62,305 (2.0) | 15,095 (0.5) | 17,895 (0.6) | 15,235 (0.5) | 17,995 (0.6) |
| | 30-39 | 153,680 (4.3) | 160,435 (4.5) | 99,185 (2.8) | 108,270 (3.0) | 24,670 (0.7) | 26,230 (0.7) | 85,920 (2.4) | 91,280 (2.5) | 16,155 (0.4) | 18,255 (0.5) | 70,220 (2.0) | 73,295 (2.0) | 16,110 (0.4) | 17,970 (0.5) | 14,355 (0.4) | 16,150 (0.4) |
| | 40-49 | 128,255 (4.0) | 133,280 (4.1) | 86,895 (2.7) | 92,995 (2.9) | 23,290 (0.7) | 24,635 (0.8) | 77,390 (2.4) | 80,990 (2.5) | 15,920 (0.5) | 17,315 (0.5) | 72,850 (2.3) | 75,325 (2.3) | 15,840 (0.5) | 17,155 (0.5) | 7,905 (0.2) | 8,595 (0.3) |
| | 50-59 | 76,600 (2.2) | 80,550 (2.3) | 47,805 (1.4) | 50,855 (1.5) | 10,960 (0.3) | 11,735 (0.3) | 46,665 (1.4) | 49,070 (1.4) | 23,330 (0.7) | 25,530 (0.7) | 46,385 (1.3) | 48,305 (1.4) | 15,690 (0.5) | 17,025 (0.5) | 7,800 (0.2) | 8,320 (0.2) |
| | 60-69 | 58,970 (2.2) | 62,460 (2.3) | 28,355 (1.0) | 30,445 (1.1) | 5,135 (0.2) | 5,555 (0.2) | 24,485 (0.9) | 26,075 (1.0) | 15,745 (0.6) | 17,165 (0.6) | 17,505 (0.6) | 18,440 (0.7) | 7,030 (0.3) | 7,685 (0.3) | 3,900 (0.1) | 4,140 (0.2) |
| | 70-79 | 31,840 (1.4) | 34,180 (1.6) | 12,275 (0.6) | 13,220 (0.6) | 1,875 (0.1) | 2,065 (0.1) | 12,300 (0.6) | 13,180 (0.6) | 6,565 (0.3) | 7,195 (0.3) | 5,920 (0.3) | 6,255 (0.3) | 2,260 (0.1) | 2,515 (0.1) | 1,475 (0.1) | 1,580 (0.1) |
| | 80+ | 15,355 (1.2) | 16,785 (1.3) | 7,425 (0.6) | 8,120 (0.6) | 1,255 (0.1) | 1,410 (0.1) | 4,635 (0.4) | 5,075 (0.4) | 7,370 (0.6) | 8,245 (0.7) | 2,400 (0.2) | 2,570 (0.2) | 1,160 (0.1) | 1,335 (0.1) | 1,230 (0.1) | 1,335 (0.1) |
| sex | Female | 326,795 (2.6) | 363,830 (2.9) | 242,560 (1.9) | 293,875 (2.3) | 57,715 (0.5) | 67,620 (0.5) | 200,910 (1.6) | 223,735 (1.8) | 59,055 (0.5) | 68,255 (0.5) | 181,750 (1.5) | 202,745 (1.6) | 50,205 (0.4) | 57,485 (0.5) | 42,970 (0.3) | 54,485 (0.4) |
| | Male | 363,630 (2.9) | 402,040 (3.2) | 257,005 (2.0) | 310,380 (2.5) | 62,895 (0.5) | 72,300 (0.6) | 196,915 (1.6) | 221,315 (1.8) | 54,520 (0.4) | 63,770 (0.5) | 187,565 (1.5) | 207,990 (1.7) | 50,675 (0.4) | 59,220 (0.5) | 39,690 (0.3) | 51,205 (0.4) |
| region | East | 108,030 (1.9) | 120,340 (2.1) | 80,525 (1.4) | 94,650 (1.6) | 36,895 (0.6) | 43,115 (0.7) | 75,155 (1.3) | 84,630 (1.5) | 23,740 (0.4) | 28,380 (0.5) | 96,865 (1.7) | 109,045 (1.9) | 27,260 (0.5) | 31,360 (0.5) | 18,975 (0.3) | 25,250 (0.4) |
| | East Midlands | 200,645 (4.6) | 224,035 (5.2) | 50,290 (1.2) | 58,690 (1.4) | 13,355 (0.3) | 15,670 (0.4) | 55,155 (1.3) | 61,890 (1.4) | 18,280 (0.4) | 21,695 (0.5) | 63,970 (1.5) | 71,285 (1.6) | 15,070 (0.3) | 17,695 (0.4) | 18,405 (0.4) | 24,435 (0.6) |
| | London | 173,335 (9.7) | 188,780 (10.5) | 51,010 (2.8) | 56,005 (3.1) | 17,725 (1.0) | 19,475 (1.1) | 126,490 (7.0) | 139,255 (7.8) | 27,865 (1.6) | 31,590 (1.8) | 74,260 (4.1) | 82,600 (4.6) | 20,960 (1.2) | 24,725 (1.4) | 9,025 (0.5) | 10,515 (0.6) |
| | North East | 8,640 (0.7) | 9,720 (0.8) | 25,385 (2.2) | 31,835 (2.7) | 2,610 (0.2) | 3,260 (0.3) | 11,860 (1.0) | 13,675 (1.2) | 2,675 (0.2) | 3,270 (0.3) | 14,750 (1.3) | 16,560 (1.4) | 2,525 (0.2) | 3,160 (0.3) | 3,445 (0.3) | 4,600 (0.4) |
| | North West | 23,000 (1.1) | 25,160 (1.2) | 21,165 (1.0) | 25,745 (1.2) | 6,705 (0.3) | 7,975 (0.4) | 17,280 (0.8) | 19,865 (0.9) | 1,135 (0.1) | 1,260 (0.1) | 13,410 (0.6) | 14,960 (0.7) | 2,925 (0.1) | 3,495 (0.2) | 1,530 (0.1) | 2,025 (0.1) |
| | South East | 21,610 (1.3) | 23,470 (1.4) | 9,420 (0.6) | 10,780 (0.7) | 7,455 (0.5) | 8,425 (0.5) | 21,190 (1.3) | 23,495 (1.4) | 2,825 (0.2) | 3,085 (0.2) | 16,005 (1.0) | 17,215 (1.0) | 4,675 (0.3) | 5,255 (0.3) | 3,385 (0.2) | 4,015 (0.2) |
| | South West | 33,820 (1.0) | 36,535 (1.0) | 4,945 (0.1) | 5,535 (0.2) | 5,095 (0.1) | 5,935 (0.2) | 27,940 (0.8) | 30,595 (0.9) | 4,815 (0.1) | 5,555 (0.2) | 15,330 (0.4) | 16,685 (0.5) | 5,590 (0.2) | 6,485 (0.2) | 7,080 (0.2) | 8,780 (0.3) |
| | West Midlands | 65,260 (6.4) | 73,425 (7.2) | 70,250 (6.9) | 77,920 (7.6) | 15,380 (1.5) | 17,375 (1.7) | 23,485 (2.3) | 25,695 (2.5) | 22,860 (2.2) | 26,040 (2.6) | 31,365 (3.1) | 33,885 (3.3) | 11,175 (1.1) | 12,265 (1.2) | 11,545 (1.1) | 13,395 (1.3) |
| | Yorkshire and The Humber | 54,790 (1.5) | 62,915 (1.7) | 185,960 (5.1) | 242,270 (6.7) | 15,340 (0.4) | 18,625 (0.5) | 38,840 (1.1) | 45,415 (1.2) | 9,200 (0.3) | 10,910 (0.3) | 42,505 (1.2) | 47,490 (1.3) | 10,490 (0.3) | 11,980 (0.3) | 8,965 (0.2) | 12,230 (0.3) |
| IMD | 1 Most deprived | 137,995 (2.8) | 152,740 (3.1) | 257,980 (5.2) | 314,575 (6.3) | 54,780 (1.1) | 63,695 (1.3) | 101,605 (2.0) | 115,695 (2.3) | 44,070 (0.9) | 51,275 (1.0) | 152,360 (3.1) | 169,330 (3.4) | 35,940 (0.7) | 41,805 (0.8) | 27,090 (0.5) | 34,760 (0.7) |
| | 2 | 210,115 (4.3) | 231,525 (4.8) | 115,185 (2.4) | 138,270 (2.8) | 31,380 (0.6) | 36,080 (0.7) | 103,580 (2.1) | 115,590 (2.4) | 27,995 (0.6) | 32,675 (0.7) | 90,415 (1.9) | 100,355 (2.1) | 24,830 (0.5) | 28,865 (0.6) | 18,200 (0.4) | 23,165 (0.5) |
| | 3 | 149,150 (2.9) | 164,265 (3.2) | 62,685 (1.2) | 73,410 (1.4) | 18,110 (0.4) | 20,705 (0.4) | 82,580 (1.6) | 91,160 (1.8) | 19,805 (0.4) | 22,805 (0.4) | 54,800 (1.1) | 60,490 (1.2) | 16,885 (0.3) | 19,395 (0.4) | 14,925 (0.3) | 18,820 (0.4) |
| | 4 | 96,690 (2.0) | 107,600 (2.2) | 33,760 (0.7) | 40,015 (0.8) | 8,920 (0.2) | 10,370 (0.2) | 58,375 (1.2) | 64,390 (1.3) | 11,930 (0.2) | 13,705 (0.3) | 35,890 (0.7) | 39,760 (0.8) | 12,065 (0.2) | 13,700 (0.3) | 11,180 (0.2) | 14,180 (0.3) |
| | 5 Least deprived | 76,390 (1.7) | 86,140 (1.9) | 20,115 (0.4) | 23,595 (0.5) | 5,095 (0.1) | 6,025 (0.1) | 40,570 (0.9) | 44,930 (1.0) | 7,100 (0.2) | 8,215 (0.2) | 22,800 (0.5) | 25,220 (0.6) | 7,905 (0.2) | 9,000 (0.2) | 8,430 (0.2) | 10,720 (0.2) |
| | Unknown | 20,090 (2.7) | 23,605 (3.2) | 9,840 (1.3) | 14,390 (1.9) | 2,320 (0.3) | 3,040 (0.4) | 11,110 (1.5) | 13,285 (1.8) | 2,670 (0.4) | 3,350 (0.5) | 13,045 (1.8) | 15,575 (2.1) | 3,255 (0.4) | 3,945 (0.5) | 2,835 (0.4) | 4,045 (0.5) |
| dementia | Present | 640 (1.4) | 695 (1.6) | 410 (0.9) | 445 (1.0) | 100 (0.2) | 115 (0.3) | 170 (0.4) | 205 (0.5) | 405 (0.9) | 460 (1.0) | 150 (0.3) | 160 (0.4) | 75 (0.2) | 80 (0.2) | 75 (0.2) | 80 (0.2) |
| | Absent | 689,785 (2.8) | 765,175 (3.1) | 499,160 (2.0) | 603,810 (2.4) | 120,510 (0.5) | 139,810 (0.6) | 397,650 (1.6) | 444,845 (1.8) | 113,170 (0.5) | 131,565 (0.5) | 369,165 (1.5) | 410,570 (1.6) | 100,805 (0.4) | 116,625 (0.5) | 82,585 (0.3) | 105,610 (0.4) |
| diabetes | Present | 118,950 (5.0) | 126,110 (5.3) | 80,610 (3.4) | 87,210 (3.6) | 22,455 (0.9) | 24,220 (1.0) | 53,210 (2.2) | 56,680 (2.4) | 23,810 (1.0) | 26,090 (1.1) | 38,955 (1.6) | 40,880 (1.7) | 10,735 (0.4) | 11,920 (0.5) | 6,155 (0.3) | 6,685 (0.3) |
| | Absent | 571,480 (2.5) | 639,760 (2.8) | 418,955 (1.8) | 517,045 (2.3) | 98,155 (0.4) | 115,705 (0.5) | 344,615 (1.5) | 388,370 (1.7) | 89,760 (0.4) | 105,935 (0.5) | 330,360 (1.5) | 369,850 (1.6) | 90,145 (0.4) | 104,785 (0.5) | 76,505 (0.3) | 99,010 (0.4) |
| hypertension | Present | 33,900 (1.8) | 36,065 (1.9) | 21,680 (1.1) | 23,480 (1.2) | 5,350 (0.3) | 5,815 (0.3) | 16,485 (0.9) | 17,580 (0.9) | 11,365 (0.6) | 12,500 (0.7) | 18,705 (1.0) | 19,620 (1.0) | 5,625 (0.3) | 6,180 (0.3) | 3,055 (0.2) | 3,280 (0.2) |
| | Absent | 656,525 (2.8) | 729,805 (3.1) | 477,885 (2.1) | 580,780 (2.5) | 115,260 (0.5) | 134,105 (0.6) | 381,340 (1.6) | 427,470 (1.8) | 102,210 (0.4) | 119,525 (0.5) | 350,610 (1.5) | 391,110 (1.7) | 95,255 (0.4) | 110,525 (0.5) | 79,605 (0.3) | 102,415 (0.4) |
| learning disability | Present | 2,210 (1.5) | 2,455 (1.7) | 4,145 (2.9) | 4,895 (3.4) | 725 (0.5) | 835 (0.6) | 1,370 (1.0) | 1,570 (1.1) | 830 (0.6) | 910 (0.6) | 1,235 (0.9) | 1,420 (1.0) | 685 (0.5) | 755 (0.5) | 625 (0.4) | 735 (0.5) |
| | Absent | 688,215 (2.8) | 763,415 (3.1) | 495,425 (2.0) | 599,360 (2.4) | 119,885 (0.5) | 139,085 (0.6) | 396,455 (1.6) | 443,480 (1.8) | 112,740 (0.5) | 131,115 (0.5) | 368,080 (1.5) | 409,315 (1.6) | 100,195 (0.4) | 115,950 (0.5) | 82,035 (0.3) | 104,960 (0.4) |

| | | White and Black African | | White and Asian | | Other Mixed | | White British | | White Irish | | Other White | | Chinese | | Any other ethnic group | |
|---|---|---|---|---|---|---|---|---|---|---|---|---|---|---|---|---|---|
| | | **SNOMED 2022** | **SNOMED 2022 with SUS data** | **SNOMED 2022** | **SNOMED 2022 with SUS data** | **SNOMED 2022** | **SNOMED 2022 with SUS data** | **SNOMED 2022** | **SNOMED 2022 with SUS data** | **SNOMED 2022** | **SNOMED 2022 with SUS data** | **SNOMED 2022** | **SNOMED 2022 with SUS data** | **SNOMED 2022** | **SNOMED 2022 with SUS data** | **SNOMED 2022** | **SNOMED 2022 with SUS data** |
| all | with records | 68,980 (0.3) | 80,555 (0.3) | 78,945 (0.3) | 99,505 (0.4) | 137,400 (0.5) | 180,605 (0.7) | 14,073,280 (56.1) | 16,948,545 (67.5) | 109,280 (0.4) | 121,220 (0.5) | 2,286,050 (9.1) | 2,496,700 (9.9) | 176,970 (0.7) | 186,855 (0.7) | 312,375 (1.2) | 394,495 (1.6) |
| age band | 0-19 | 24,225 (0.4) | 33,590 (0.6) | 31,610 (0.6) | 48,215 (0.9) | 50,585 (0.9) | 81,100 (1.5) | 2,364,790 (42.9) | 3,441,095 (62.5) | 8,280 (0.2) | 11,695 (0.2) | 381,250 (6.9) | 498,520 (9.0) | 24,620 (0.4) | 29,300 (0.5) | 68,940 (1.3) | 109,505 (2.0) |
| | 20-29 | 10,740 (0.3) | 11,550 (0.4) | 14,230 (0.5) | 15,730 (0.5) | 23,005 (0.7) | 27,060 (0.9) | 1,417,355 (45.3) | 1,797,380 (57.5) | 11,260 (0.4) | 12,295 (0.4) | 347,270 (11.1) | 370,735 (11.9) | 61,625 (2.0) | 62,780 (2.0) | 53,355 (1.7) | 66,405 (2.1) |
| | 30-39 | 11,510 (0.3) | 11,990 (0.3) | 12,810 (0.4) | 13,770 (0.4) | 25,720 (0.7) | 28,840 (0.8) | 1,772,795 (49.3) | 2,041,980 (56.8) | 16,765 (0.5) | 17,705 (0.5) | 560,950 (15.6) | 584,915 (16.3) | 33,710 (0.9) | 34,775 (1.0) | 70,895 (2.0) | 81,630 (2.3) |
| | 40-49 | 11,125 (0.3) | 11,510 (0.4) | 9,290 (0.3) | 9,830 (0.3) | 18,155 (0.6) | 20,045 (0.6) | 1,749,745 (54.2) | 1,960,865 (60.7) | 15,540 (0.5) | 16,340 (0.5) | 415,770 (12.9) | 430,825 (13.3) | 24,945 (0.8) | 25,800 (0.8) | 57,160 (1.8) | 63,590 (2.0) |
| | 50-59 | 6,840 (0.2) | 7,130 (0.2) | 6,140 (0.2) | 6,560 (0.2) | 11,115 (0.3) | 12,690 (0.4) | 2,266,255 (65.9) | 2,540,685 (73.8) | 17,620 (0.5) | 18,925 (0.6) | 255,685 (7.4) | 267,650 (7.8) | 16,005 (0.5) | 16,745 (0.5) | 32,830 (1.0) | 37,525 (1.1) |
| | 60-69 | 2,980 (0.1) | 3,140 (0.1) | 2,930 (0.1) | 3,205 (0.1) | 5,555 (0.2) | 6,515 (0.2) | 1,952,110 (71.2) | 2,188,565 (79.8) | 15,285 (0.6) | 16,515 (0.6) | 159,575 (5.8) | 167,970 (6.1) | 9,935 (0.4) | 10,630 (0.4) | 17,265 (0.6) | 20,605 (0.8) |
| | 70-79 | 1,160 (0.1) | 1,220 (0.1) | 1,375 (0.1) | 1,550 (0.1) | 2,280 (0.1) | 2,935 (0.1) | 1,663,155 (75.5) | 1,889,605 (85.8) | 15,065 (0.7) | 16,635 (0.8) | 105,165 (4.8) | 111,065 (5.0) | 4,385 (0.2) | 4,850 (0.2) | 8,400 (0.4) | 10,440 (0.5) |
| | 80+ | 400 (0.0) | 425 (0.0) | 560 (0.0) | 650 (0.1) | 975 (0.1) | 1,415 (0.1) | 887,065 (70.7) | 1,088,360 (86.7) | 9,465 (0.8) | 11,105 (0.9) | 60,385 (4.8) | 65,015 (5.2) | 1,740 (0.1) | 1,975 (0.2) | 3,530 (0.3) | 4,795 (0.4) |
| sex | Female | 34,340 (0.3) | 40,070 (0.3) | 40,000 (0.3) | 50,140 (0.4) | 69,370 (0.6) | 89,685 (0.7) | 7,252,300 (57.9) | 8,666,365 (69.1) | 54,965 (0.4) | 61,080 (0.5) | 1,153,810 (9.2) | 1,260,105 (10.1) | 92,015 (0.7) | 97,325 (0.8) | 147,170 (1.2) | 186,080 (1.5) |
| | Male | 34,640 (0.3) | 40,485 (0.3) | 38,945 (0.3) | 49,365 (0.4) | 68,025 (0.5) | 90,920 (0.7) | 6,820,980 (54.3) | 8,282,180 (65.9) | 54,315 (0.4) | 60,140 (0.5) | 1,132,240 (9.0) | 1,236,590 (9.8) | 84,955 (0.7) | 89,530 (0.7) | 165,205 (1.3) | 208,420 (1.7) |
| region | East | 20,295 (0.3) | 24,090 (0.4) | 17,785 (0.3) | 23,130 (0.4) | 32,775 (0.6) | 43,720 (0.8) | 3,207,190 (55.2) | 3,901,430 (67.2) | 28,660 (0.5) | 32,650 (0.6) | 620,700 (10.7) | 683,255 (11.8) | 38,980 (0.7) | 41,610 (0.7) | 55,075 (0.9) | 68,805 (1.2) |
| | East Midlands | 10,760 (0.2) | 12,700 (0.3) | 13,030 (0.3) | 16,530 (0.4) | 18,795 (0.4) | 28,235 (0.7) | 2,491,890 (57.4) | 2,976,340 (68.5) | 14,410 (0.3) | 16,095 (0.4) | 361,140 (8.3) | 394,435 (9.1) | 24,830 (0.6) | 26,205 (0.6) | 43,050 (1.0) | 54,200 (1.2) |
| | London | 8,735 (0.5) | 9,735 (0.5) | 11,270 (0.6) | 13,110 (0.7) | 31,965 (1.8) | 37,475 (2.1) | 348,760 (19.4) | 392,040 (21.8) | 22,180 (1.2) | 23,850 (1.3) | 404,150 (22.5) | 437,875 (24.4) | 42,690 (2.4) | 44,290 (2.5) | 90,345 (5.0) | 117,735 (6.6) |
| | North East | 2,750 (0.2) | 3,305 (0.3) | 3,110 (0.3) | 4,175 (0.4) | 4,160 (0.4) | 6,095 (0.5) | 742,000 (63.3) | 902,025 (77.0) | 2,655 (0.2) | 3,020 (0.3) | 66,585 (5.7) | 72,405 (6.2) | 6,545 (0.6) | 7,065 (0.6) | 14,395 (1.2) | 19,430 (1.7) |
| | North West | 2,980 (0.1) | 3,675 (0.2) | 3,995 (0.2) | 5,380 (0.2) | 5,920 (0.3) | 7,985 (0.4) | 1,462,975 (67.8) | 1,769,575 (82.0) | 5,300 (0.2) | 5,835 (0.3) | 101,985 (4.7) | 107,465 (5.0) | 9,635 (0.4) | 10,390 (0.5) | 13,900 (0.6) | 18,975 (0.9) |
| | South East | 4,635 (0.3) | 5,365 (0.3) | 5,805 (0.4) | 6,805 (0.4) | 9,890 (0.6) | 12,170 (0.7) | 963,370 (58.6) | 1,169,575 (71.1) | 7,740 (0.5) | 8,565 (0.5) | 157,435 (9.6) | 170,525 (10.4) | 10,125 (0.6) | 10,660 (0.6) | 20,345 (1.2) | 23,400 (1.4) |
| | South West | 5,790 (0.2) | 6,845 (0.2) | 8,515 (0.2) | 10,420 (0.3) | 12,570 (0.4) | 16,925 (0.5) | 2,256,880 (64.8) | 2,729,835 (78.4) | 11,385 (0.3) | 12,675 (0.4) | 261,570 (7.5) | 284,000 (8.2) | 15,010 (0.4) | 15,935 (0.5) | 21,635 (0.6) | 26,855 (0.8) |
| | West Midlands | 3,830 (0.4) | 4,160 (0.4) | 5,390 (0.5) | 6,060 (0.6) | 8,460 (0.8) | 9,820 (1.0) | 461,205 (45.3) | 530,980 (52.1) | 6,655 (0.7) | 7,150 (0.7) | 72,325 (7.1) | 77,690 (7.6) | 10,710 (1.1) | 11,205 (1.1) | 17,345 (1.7) | 20,605 (2.0) |
| | Yorkshire and The Humber | 8,900 (0.2) | 10,325 (0.3) | 9,895 (0.3) | 13,665 (0.4) | 12,675 (0.3) | 17,850 (0.5) | 2,118,540 (58.2) | 2,552,035 (70.1) | 10,160 (0.3) | 11,235 (0.3) | 236,545 (6.5) | 264,945 (7.3) | 18,300 (0.5) | 19,335 (0.5) | 35,835 (1.0) | 43,900 (1.2) |
| IMD | 1 Most deprived | 22,300 (0.4) | 25,555 (0.5) | 17,080 (0.3) | 21,885 (0.4) | 33,620 (0.7) | 44,835 (0.9) | 2,329,005 (46.9) | 2,848,830 (57.3) | 17,825 (0.4) | 19,750 (0.4) | 535,420 (10.8) | 584,140 (11.8) | 24,690 (0.5) | 26,285 (0.5) | 93,600 (1.9) | 117,880 (2.4) |
| | 2 | 15,330 (0.3) | 17,890 (0.4) | 15,285 (0.3) | 19,035 (0.4) | 30,380 (0.6) | 39,585 (0.8) | 2,494,495 (51.2) | 3,015,755 (61.9) | 21,645 (0.4) | 23,995 (0.5) | 513,060 (10.5) | 560,910 (11.5) | 36,285 (0.7) | 38,145 (0.8) | 73,625 (1.5) | 93,370 (1.9) |
| | 3 | 11,945 (0.2) | 13,955 (0.3) | 15,340 (0.3) | 18,885 (0.4) | 26,655 (0.5) | 34,455 (0.7) | 2,960,575 (58.0) | 3,531,960 (69.2) | 24,175 (0.5) | 26,685 (0.5) | 475,990 (9.3) | 517,505 (10.1) | 36,180 (0.7) | 37,995 (0.7) | 57,515 (1.1) | 72,380 (1.4) |
| | 4 | 9,725 (0.2) | 11,415 (0.2) | 14,795 (0.3) | 18,500 (0.4) | 23,630 (0.5) | 30,635 (0.6) | 3,039,835 (61.5) | 3,629,195 (73.4) | 23,555 (0.5) | 25,985 (0.5) | 405,530 (8.2) | 441,190 (8.9) | 37,465 (0.8) | 39,465 (0.8) | 47,955 (1.0) | 59,660 (1.2) |
| | 5 Least deprived | 7,080 (0.2) | 8,425 (0.2) | 13,910 (0.3) | 17,585 (0.4) | 18,775 (0.4) | 24,440 (0.5) | 2,868,285 (64.0) | 3,427,485 (76.5) | 19,650 (0.4) | 21,920 (0.5) | 302,375 (6.7) | 328,395 (7.3) | 33,070 (0.7) | 35,100 (0.8) | 30,985 (0.7) | 39,325 (0.9) |
| | Unknown | 2,605 (0.4) | 3,310 (0.4) | 2,530 (0.3) | 3,610 (0.5) | 4,340 (0.6) | 6,655 (0.9) | 381,085 (51.5) | 495,315 (67.0) | 2,430 (0.3) | 2,890 (0.4) | 53,675 (7.3) | 64,560 (8.7) | 9,285 (1.3) | 9,865 (1.3) | 8,695 (1.2) | 11,885 (1.6) |
| dementia | Present | 20 (0.0) | 20 (0.0) | 25 (0.1) | 25 (0.1) | 35 (0.1) | 50 (0.1) | 31,845 (71.5) | 38,750 (87.0) | 405 (0.9) | 470 (1.1) | 2,110 (4.7) | 2,265 (5.1) | 45 (0.1) | 65 (0.1) | 130 (0.3) | 180 (0.4) |
| | Absent | 68,960 (0.3) | 80,535 (0.3) | 78,920 (0.3) | 99,480 (0.4) | 137,360 (0.5) | 180,555 (0.7) | 14,041,435 (56.0) | 16,909,795 (67.5) | 108,875 (0.4) | 120,750 (0.5) | 2,283,940 (9.1) | 2,494,435 (10.0) | 176,925 (0.7) | 186,795 (0.7) | 312,245 (1.2) | 394,315 (1.6) |
| diabetes | Present | 5,020 (0.2) | 5,310 (0.2) | 4,750 (0.2) | 5,245 (0.2) | 7,985 (0.3) | 9,225 (0.4) | 1,488,060 (62.1) | 1,731,290 (72.3) | 12,845 (0.5) | 14,215 (0.6) | 142,215 (5.9) | 150,625 (6.3) | 8,935 (0.4) | 9,765 (0.4) | 24,810 (1.0) | 29,635 (1.2) |
| | Absent | 63,960 (0.3) | 75,245 (0.3) | 74,195 (0.3) | 94,260 (0.4) | 129,415 (0.6) | 171,380 (0.8) | 12,585,220 (55.4) | 15,217,255 (67.0) | 96,435 (0.4) | 107,005 (0.5) | 2,143,835 (9.4) | 2,346,070 (10.3) | 168,040 (0.7) | 177,095 (0.8) | 287,570 (1.3) | 364,860 (1.6) |
| hypertension | Present | 2,725 (0.1) | 2,865 (0.2) | 1,865 (0.1) | 2,045 (0.1) | 3,495 (0.2) | 4,225 (0.2) | 1,367,220 (72.5) | 1,561,030 (82.8) | 10,010 (0.5) | 11,095 (0.6) | 106,360 (5.6) | 112,005 (5.9) | 4,240 (0.2) | 4,615 (0.2) | 9,295 (0.5) | 11,470 (0.6) |
| | Absent | 66,250 (0.3) | 77,685 (0.3) | 77,075 (0.3) | 97,460 (0.4) | 133,905 (0.6) | 176,380 (0.8) | 12,706,065 (54.7) | 15,387,515 (66.3) | 99,270 (0.4) | 110,125 (0.5) | 2,179,690 (9.4) | 2,384,690 (10.3) | 172,730 (0.7) | 182,245 (0.8) | 303,080 (1.3) | 383,030 (1.6) |
| learning disability | Present | 295 (0.2) | 350 (0.2) | 355 (0.2) | 445 (0.3) | 725 (0.5) | 920 (0.6) | 99,135 (69.3) | 113,130 (79.1) | 405 (0.3) | 440 (0.3) | 6,610 (4.6) | 7,145 (5.0) | 215 (0.2) | 240 (0.2) | 825 (0.6) | 1,140 (0.8) |
| | Absent | 68,685 (0.3) | 80,205 (0.3) | 78,590 (0.3) | 99,060 (0.4) | 136,675 (0.5) | 179,685 (0.7) | 13,974,145 (56.0) | 16,835,410 (67.5) | 108,875 (0.4) | 120,780 (0.5) | 2,279,440 (9.1) | 2,489,550 (10.0) | 176,760 (0.7) | 186,615 (0.7) | 311,550 (1.2) | 393,355 (1.6) |
