## Extended Data Table 4 for "Consistency, completeness and external validity of ethnicity recording in NHS primary care records: a cohort study in 25 million patients’ records at source using OpenSAFELY"

Table 4: Count of patients' most frequently recorded ethnicity (proportion of latest ethnicity).

| Latest Recorded Ethnicity | Most Frequent Ethnicity | | | | |
|---|---|---|---|---|---|
| | Asian | Black | Mixed | White | Other |
| Asian | 1,695,745 (99.3) | 1,165 (0.1) | 1,995 (0.1) | 4,690 (0.3) | 4,835 (0.3) |
| Black | 4,380 (0.8) | 569,290 (97.5) | 3,825 (0.7) | 5,450 (0.9) | 825 (0.1) |
| Mixed | 15,155 (4.1) | 26,300 (7.1) | 309,220 (84.0) | 15,080 (4.1) | 2,225 (0.6) |
| White | 23,975 (0.1) | 16,845 (0.1) | 32,920 (0.2) | 16,390,425 (99.5) | 4,445 (0.0) |
| Other | 23,970 (4.9) | 6,825 (1.4) | 9,520 (1.9) | 49,600 (10.1) | 399,440 (81.6) |
