## Extended Data Table 6 for "Consistency, completeness and external validity of ethnicity recording in NHS primary care records: a cohort study in 25 million patients’ records at source using OpenSAFELY"

Table 6: Count of patients with a recorded ethnicity in Secondary Care by ethnicity group excluding Unknown ethnicites (proportion of Primary Care population). All counts are rounded to the nearest 5.

| Primary Care ethnicity | Secondary Care ethnicity | | | | |
|---|---|---|---|---|---|
| | **Asian** | **Black** | **Mixed** | **White** | **Other** |
| Asian (1,708,430) | 822,555 (48.1) | 6,935 (0.4) | 23,025 (1.3) | 37,000 (2.2) | 85,035 (5) |
| Black (583,770) | 9,080 (1.6) | 249,215 (42.7) | 23,625 (4) | 22,660 (3.9) | 27,875 (4.8) |
| Mixed (367,980) | 19,015 (5.2) | 34,550 (9.4) | 75,460 (20.5) | 62,030 (16.9) | 25,640 (7) |
| White (16,468,610) | 39,165 (0.2) | 29,200 (0.2) | 105,610 (0.6) | 10,871,855 (66) | 173,795 (1.1) |
| Other (489,350) | 32,475 (6.6) | 6,980 (1.4) | 12,575 (2.6) | 59,635 (12.2) | 91,125 (18.6) |
