## Extended Data Table 7 for "Consistency, completeness and external validity of ethnicity recording in NHS primary care records: a cohort study in 25 million patients’ records at source using OpenSAFELY"

Table 7: Count of patients with a recorded ethnicity in OpenSAFELY TPP by ethnicity group (proportion of registered TPP population) and 2021 ONS Census counts [amended to 2001 grouping] (proportion of 2021 ONS Census population). All counts are rounded to the nearest 5.

| | Asian | | | Black | | | Mixed | | | White | | | Other | | |
|---|---|---|---|---|---|---|---|---|---|---|---|---|---|---|---|
| | **SNOMED 2022** | **SNOMED 2022 with SUS data** | **2021 ONS Census [amended to 2001 grouping]** | **SNOMED 2022** | **SNOMED 2022 with SUS data** | **2021 ONS Census [amended to 2001 grouping]** | **SNOMED 2022** | **SNOMED 2022 with SUS data** | **2021 ONS Census [amended to 2001 grouping]** | **SNOMED 2022** | **SNOMED 2022 with SUS data** | **2021 ONS Census [amended to 2001 grouping]** | **SNOMED 2022** | **SNOMED 2022 with SUS data** | **2021 ONS Census [amended to 2001 grouping]** |
| England | 1,708,430 (8.71) | 1,955,095 (8.42) | 4,995,225 (8.84) | 583,770 (2.98) | 659,410 (2.84) | 2,381,720 (4.22) | 367,980 (1.88) | 466,220 (2.01) | 1,669,380 (2.96) | 16,468,610 (83.95) | 19,566,635 (84.23) | 45,783,400 (81.05) | 489,350 (2.49) | 581,395 (2.5) | 1,660,315 (2.94) |
| East Midlands | 319,445 (9.36) | 360,305 (8.96) | 368,130 (7.54) | 97,320 (2.85) | 110,695 (2.75) | 129,985 (2.66) | 60,985 (1.79) | 81,860 (2.04) | 117,245 (2.4) | 2,867,445 (84.01) | 3,386,935 (84.25) | 4,179,775 (85.65) | 67,880 (1.99) | 80,350 (2) | 84,915 (1.74) |
| East of England | 300,605 (6.7) | 342,735 (6.4) | 367,425 (5.8) | 147,865 (3.29) | 168,770 (3.15) | 184,950 (2.92) | 89,825 (2) | 116,180 (2.17) | 179,655 (2.84) | 3,856,555 (85.91) | 4,617,355 (86.22) | 5,478,365 (86.48) | 94,055 (2.1) | 110,425 (2.06) | 124,675 (1.97) |
| London | 368,555 (25.23) | 403,495 (24.77) | 1,670,120 (18.98) | 123,080 (8.43) | 138,875 (8.53) | 1,188,370 (13.5) | 60,995 (4.18) | 70,810 (4.35) | 505,775 (5.75) | 775,090 (53.06) | 853,735 (52.41) | 4,731,170 (53.76) | 133,035 (9.11) | 162,100 (9.95) | 704,290 (8) |
| North East | 48,500 (5.31) | 58,495 (5.3) | 83,605 (3.16) | 19,950 (2.18) | 22,990 (2.08) | 26,635 (1.01) | 13,465 (1.47) | 18,160 (1.65) | 33,270 (1.26) | 811,235 (88.75) | 977,415 (88.57) | 2,462,720 (93.04) | 20,940 (2.29) | 26,485 (2.4) | 40,785 (1.54) |
| North West | 68,150 (4.02) | 78,725 (3.88) | 568,635 (7.67) | 17,470 (1.03) | 19,705 (0.97) | 173,920 (2.34) | 14,420 (0.85) | 19,055 (0.94) | 163,245 (2.2) | 1,570,265 (92.7) | 1,882,895 (92.77) | 6,347,395 (85.57) | 23,535 (1.39) | 29,350 (1.45) | 164,205 (2.21) |
| South East | 59,670 (4.71) | 66,180 (4.4) | 586,215 (6.32) | 23,510 (1.86) | 25,550 (1.7) | 221,585 (2.39) | 23,715 (1.87) | 28,355 (1.89) | 260,870 (2.81) | 1,128,545 (89.15) | 1,348,710 (89.74) | 8,009,380 (86.33) | 30,470 (2.41) | 34,070 (2.27) | 200,010 (2.16) |
| South West | 71,795 (2.66) | 78,600 (2.44) | 132,670 (2.33) | 25,735 (0.95) | 28,730 (0.89) | 69,615 (1.22) | 33,950 (1.26) | 42,945 (1.33) | 114,075 (2) | 2,529,835 (93.77) | 3,026,520 (94) | 5,309,610 (93.13) | 36,640 (1.36) | 42,795 (1.33) | 75,220 (1.32) |
| West Midlands | 174,370 (20.83) | 194,405 (20.51) | 760,965 (12.79) | 65,400 (7.81) | 72,195 (7.62) | 269,020 (4.52) | 29,220 (3.49) | 33,420 (3.53) | 178,225 (2.99) | 540,185 (64.52) | 615,885 (64.99) | 4,585,025 (77.05) | 28,055 (3.35) | 31,815 (3.36) | 157,525 (2.65) |
| Yorkshire and the Humber | 294,930 (10.47) | 369,235 (10.91) | 457,465 (8.35) | 62,195 (2.21) | 70,370 (2.08) | 117,645 (2.15) | 40,435 (1.44) | 54,080 (1.6) | 117,015 (2.14) | 2,365,245 (83.97) | 2,828,245 (83.55) | 4,679,965 (85.39) | 54,135 (1.92) | 63,245 (1.87) | 108,685 (1.98) |
