## Extended Data Table 8 for "Consistency, completeness and external validity of ethnicity recording in NHS primary care records: a cohort study in 25 million patients’ records at source using OpenSAFELY"

Table 8: Count of patients with a recorded ethnicity in OpenSAFELY TPP [amended to the 2021 ethnicity grouping] (proportion of registered TPP population) and 2021 ONS Census counts (proportion of 2021 ONS Census population). All counts are rounded to the nearest 5.

| | Asian | | | Black | | | Mixed | | | White | | | Other | | |
|---|---|---|---|---|---|---|---|---|---|---|---|---|---|---|---|
| | **SNOMED 2022 (amended to 2021 grouping)** | **SNOMED 2022 with SUS data (amended to 2021 grouping)** | **2021 ONS Census** | **SNOMED 2022 (amended to 2021 grouping)** | **SNOMED 2022 with SUS data (amended to 2021 grouping)** | **2021 ONS Census** | **SNOMED 2022 (amended to 2021 grouping)** | **SNOMED 2022 with SUS data (amended to 2021 grouping)** | **2021 ONS Census** | **SNOMED 2022 (amended to 2021 grouping)** | **SNOMED 2022 with SUS data (amended to 2021 grouping)** | **2021 ONS Census** | **SNOMED 2022 (amended to 2021 grouping)** | **SNOMED 2022 with SUS data (amended to 2021 grouping)** | **2021 ONS Census** |
| England | 1,885,400 (9.61) | 2,141,950 (9.22) | 5,426,390 (9.61) | 583,770 (2.98) | 659,460 (2.84) | 2,381,720 (4.22) | 367,985 (1.88) | 466,360 (2.01) | 1,669,380 (2.96) | 16,468,610 (83.95) | 19,566,465 (84.23) | 45,783,400 (81.05) | 312,375 (1.59) | 394,495 (1.7) | 1,229,150 (2.18) |
| East Midlands | 344,275 (10.09) | 386,490 (9.61) | 391,105 (8.01) | 97,320 (2.85) | 110,675 (2.75) | 129,985 (2.66) | 60,990 (1.79) | 81,900 (2.04) | 117,245 (2.4) | 2,867,440 (84.01) | 3,386,870 (84.25) | 4,179,775 (85.65) | 43,050 (1.26) | 54,200 (1.35) | 61,945 (1.27) |
| East of England | 339,585 (7.56) | 384,345 (7.18) | 405,870 (6.41) | 147,865 (3.29) | 168,785 (3.15) | 184,950 (2.92) | 89,830 (2) | 116,190 (2.17) | 179,655 (2.84) | 3,856,550 (85.91) | 4,617,335 (86.22) | 5,478,365 (86.48) | 55,075 (1.23) | 68,805 (1.28) | 86,230 (1.36) |
| London | 411,250 (28.15) | 447,805 (27.49) | 1,817,640 (20.66) | 123,085 (8.43) | 138,915 (8.53) | 1,188,370 (13.5) | 60,995 (4.18) | 70,835 (4.35) | 505,775 (5.75) | 775,090 (53.06) | 853,765 (52.41) | 4,731,170 (53.76) | 90,345 (6.18) | 117,735 (7.23) | 556,770 (6.33) |
| North East | 55,040 (6.02) | 65,555 (5.94) | 98,045 (3.7) | 19,950 (2.18) | 22,990 (2.08) | 26,635 (1.01) | 13,465 (1.47) | 18,175 (1.65) | 33,270 (1.26) | 811,240 (88.75) | 977,450 (88.57) | 2,462,720 (93.04) | 14,395 (1.57) | 19,430 (1.76) | 26,340 (1) |
| North West | 77,785 (4.59) | 89,135 (4.39) | 622,685 (8.39) | 17,470 (1.03) | 19,715 (0.97) | 173,920 (2.34) | 14,425 (0.85) | 19,065 (0.94) | 163,245 (2.2) | 1,570,260 (92.7) | 1,882,875 (92.76) | 6,347,395 (85.57) | 13,900 (0.82) | 18,975 (0.93) | 110,155 (1.49) |
| South East | 69,800 (5.51) | 76,830 (5.11) | 650,545 (7.01) | 23,505 (1.86) | 25,555 (1.7) | 221,585 (2.39) | 23,715 (1.87) | 28,355 (1.89) | 260,870 (2.81) | 1,128,545 (89.15) | 1,348,665 (89.74) | 8,009,380 (86.33) | 20,345 (1.61) | 23,400 (1.56) | 135,685 (1.46) |
| South West | 86,810 (3.22) | 94,535 (2.94) | 159,185 (2.79) | 25,735 (0.95) | 28,725 (0.89) | 69,615 (1.22) | 33,955 (1.26) | 42,970 (1.33) | 114,075 (2) | 2,529,835 (93.77) | 3,026,510 (94) | 5,309,610 (93.13) | 21,635 (0.8) | 26,855 (0.83) | 48,705 (0.85) |
| West Midlands | 185,085 (22.11) | 205,620 (21.7) | 794,265 (13.35) | 65,400 (7.81) | 72,190 (7.62) | 269,020 (4.52) | 29,225 (3.49) | 33,435 (3.53) | 178,225 (2.99) | 540,185 (64.52) | 615,820 (64.98) | 4,585,025 (77.05) | 17,345 (2.07) | 20,605 (2.17) | 124,225 (2.09) |
| Yorkshire and the Humber | 313,230 (11.12) | 388,560 (11.48) | 487,055 (8.89) | 62,195 (2.21) | 70,380 (2.08) | 117,645 (2.15) | 40,435 (1.44) | 54,070 (1.6) | 117,015 (2.14) | 2,365,245 (83.97) | 2,828,215 (83.55) | 4,679,965 (85.39) | 35,835 (1.27) | 43,900 (1.3) | 79,095 (1.44) |
