## Extended Data Table 9 for "Consistency, completeness and external validity of ethnicity recording in NHS primary care records: a cohort study in 25 million patients’ records at source using OpenSAFELY"

Table 9: Count of individual ethnicity code use

| Code | Term | Count |
|---|---|---|
| 92491000000104 | African - ethnic category 2001 census | 369350 |
| 978231000000100 | African: African, African Scottish or African British - Scotland ethnic category 2011 census | 535 |
| 978251000000107 | African: any other African - Scotland ethnic category 2011 census | 195 |
| 413465009 | Afro-Caribbean | 1435 |
| 413466005 | Afro-Caucasian | 90 |
| 88971000000106 | Albanian - ethnic category 2001 census | 10245 |
| 94151000000105 | Any other group - ethnic category 2001 census | 44060 |
| 89001000000105 | Arab - ethnic category 2001 census | 53120 |
| 90027003 | Arabs | 2990 |
| 315280000 | Asian - ethnic group | 81595 |
| 92611000000106 | Asian and Chinese - ethnic category 2001 census | 5680 |
| 976831000000100 | Asian or Asian British: Bangladeshi - England and Wales ethnic category 2011 census | 4605 |
| 977731000000108 | Asian or Asian British: Bangladeshi - Northern Ireland ethnic category 2011 census | 600 |
| 976851000000107 | Asian or Asian British: Chinese - England and Wales ethnic category 2011 census | 5405 |
| 977751000000101 | Asian or Asian British: Chinese - Northern Ireland ethnic category 2011 census | 260 |
| 976791000000107 | Asian or Asian British: Indian - England and Wales ethnic category 2011 census | 32355 |
| 977591000000103 | Asian or Asian British: Indian - Northern Ireland ethnic category 2011 census | 5975 |
| 976811000000108 | Asian or Asian British: Pakistani - England and Wales ethnic category 2011 census | 17915 |
| 977711000000100 | Asian or Asian British: Pakistani - Northern Ireland ethnic category 2011 census | 2790 |
| 976871000000103 | Asian or Asian British: any other Asian background - England and Wales ethnic category 2011 census | 13365 |
| 977771000000105 | Asian or Asian British: any other Asian background - Northern Ireland ethnic category 2011 census | 1110 |
| 978171000000105 | Asian or Asian Scottish or Asian British: Bangladeshi, Bangladeshi Scottish or Bangladeshi British - Scotland ethnic category 2011 census | 185 |
| 978191000000109 | Asian or Asian Scottish or Asian British: Chinese - Scotland ethnic category 2011 census | 60 |
| 978111000000100 | Asian or Asian Scottish or Asian British: Indian, Indian Scottish or Indian British - Scotland ethnic category 2011 census | 1545 |
| 978071000000106 | Asian or Asian Scottish or Asian British: Pakistani, Pakistani Scottish or Pakistani British - Scotland ethnic category 2011 census | 1080 |
| 978211000000108 | Asian or Asian Scottish or Asian British: any other Asian group - Scotland ethnic category 2011 census | 310 |
| 88951000000102 | Baltic States (Estonian or Latvian or Lithuanian) - ethnic category 2001 census | 86330 |
| 186003008 | Bangladeshi | 29650 |

Table 9: Count of individual ethnicity code use

| | | |
|---|---|---|
| 92471000000103 | Bangladeshi or British Bangladeshi - ethnic category 2001 census | 124530 |
| 315240009 | Black - ethnic group | 16180 |
| 185993005 | Black - other African country | 1425 |
| 185996002 | Black - other Asian | 820 |
| 185998001 | Black - other, mixed | 4230 |
| 18167009 | Black African | 135180 |
| 315635008 | Black African and White | 8365 |
| 275587000 | Black Arab | 455 |
| 185990008 | Black British | 17280 |
| 110791000000100 | Black British - ethnic category 2001 census | 31880 |
| 185988007 | Black Caribbean | 32445 |
| 315634007 | Black Caribbean and White | 12520 |
| 270460000 | Black Caribbean/West India/Guyana | 4155 |
| 275589002 | Black East African Asian | 185 |
| 270462008 | Black East African Asian/Indo-Caribbean | 255 |
| 309644006 | Black Guyana | 105 |
| 185995003 | Black Indian sub-continent | 310 |
| 275590006 | Black Indo-Caribbean | 55 |
| 275588005 | Black Iranian | 100 |
| 270461001 | Black N African/Arab/Iranian | 630 |
| 275586009 | Black North African | 270 |
| 309643000 | Black West Indian | 600 |
| 92581000000100 | Black and Asian - ethnic category 2001 census | 1210 |
| 92591000000103 | Black and Chinese - ethnic category 2001 census | 115 |
| 110771000000104 | Black and White - ethnic category 2001 census | 3000 |
| 976891000000104 | Black or African or Caribbean or Black British: African - England and Wales ethnic category 2011 census | 12425 |
| 977791000000109 | Black or African or Caribbean or Black British: African - Northern Ireland ethnic category 2011 census | 620 |
| 976911000000101 | Black or African or Caribbean or Black British: Caribbean - England and Wales ethnic category 2011 census | 5790 |
| 977811000000105 | Black or African or Caribbean or Black British: Caribbean - Northern Ireland ethnic category 2011 census | 315 |
| 976931000000109 | Black or African or Caribbean or Black British: other Black or African or Caribbean background - England and Wales ethnic | 3870 |

Table 9: Count of individual ethnicity code use

| | | |
|---|---|---|
| | category 2011 census | |
| 977831000000102 | Black or African or Caribbean or Black British: other Black or African or Caribbean background - Northern Ireland ethnic category 2011 census | 600 |
| 185989004 | Black, other, non-mixed origin | 2750 |
| 93991000000103 | Bosnian - ethnic category 2001 census | 800 |
| 92681000000104 | British Asian - ethnic category 2001 census | 26100 |
| 186006000 | British ethnic minority specified (NMO) | 430 |
| 186007009 | British ethnic minority unspecified (NMO) | 665 |
| 92391000000108 | British or mixed British - ethnic category 2001 census | 9407055 |
| 29343004 | Bulgarian | 11685 |
| 718962008 | Bulgarian Roma | 115 |
| 107691000000105 | Caribbean - ethnic category 2001 census | 112475 |
| 92691000000102 | Caribbean Asian - ethnic category 2001 census | 535 |
| 270463003 | Caribbean I./W.I./Guyana (NMO) | 155 |
| 275591005 | Caribbean Island (NMO) | 280 |
| 978341000000102 | Caribbean or Black: Black, Black Scottish or Black British - Scotland ethnic category 2011 census | 110 |
| 978271000000103 | Caribbean or Black: Caribbean, Caribbean Scottish or Caribbean British - Scotland ethnic category 2011 census | 145 |
| 978361000000101 | Caribbean or Black: any other Black or Caribbean group - Scotland ethnic category 2011 census | 110 |
| 413773004 | Caucasian (racial group) | 12335 |
| 33897005 | Chinese | 48705 |
| 92511000000107 | Chinese - ethnic category 2001 census | 197525 |
| 92601000000109 | Chinese and White - ethnic category 2001 census | 2445 |
| 88961000000104 | Commonwealth of (Russian) Independent States - ethnic category 2001 census | 7325 |
| 186040000 | Cook Island Maori | 25 |
| 92571000000102 | Cornish - ethnic category 2001 census | 10390 |
| 94001000000108 | Croatian - ethnic category 2001 census | 1525 |
| 92791000000109 | Cypriot (part not stated) - ethnic category 2001 census | 1645 |
| 286009 | Czech | 7000 |
| 718959005 | Czech Roma | 115 |
| 270465005 | E Afric Asian/Indo-Carib (NMO) | 105 |
| 275596000 | East African Asian (NMO) | 295 |

Table 9: Count of individual ethnicity code use

| | | |
|---|---|---|
| 92661000000108 | East African Asian - ethnic category 2001 census | 1840 |
| 110761000000106 | English - ethnic category 2001 census | 387530 |
| 69865008 | Fijian | 520 |
| 92771000000105 | Filipino - ethnic category 2001 census | 16010 |
| 275599007 | Greek (NMO) | 1570 |
| 93931000000104 | Greek - ethnic category 2001 census | 11685 |
| 275600005 | Greek Cypriot (NMO) | 400 |
| 93941000000108 | Greek Cypriot - ethnic category 2001 census | 2590 |
| 270466006 | Greek/Greek Cypriot (NMO) | 480 |
| 275593008 | Guyana (NMO) | 95 |
| 40182006 | Gypsies | 530 |
| 88931000000109 | Gypsy/Romany - ethnic category 2001 census | 12435 |
| 718963003 | Hungarian Roma | 3675 |
| 414481008 | Indian (racial group) | 203675 |
| 110751000000108 | Indian or British Indian - ethnic category 2001 census | 757205 |
| 186012005 | Indian sub-continent (NMO) | 825 |
| 275597009 | Indo-Caribbean (NMO) | 80 |
| 275595001 | Iranian (NMO) | 2565 |
| 89011000000107 | Iranian - ethnic category 2001 census | 19215 |
| 186014006 | Irish (NMO) | 705 |
| 92401000000106 | Irish - ethnic category 2001 census | 92830 |
| 977371000000109 | Irish Traveller - Northern Ireland ethnic category 2011 census | 10 |
| 88911000000101 | Irish Traveller - ethnic category 2001 census | 815 |
| 315283003 | Irish traveller | 420 |
| 94081000000103 | Israeli - ethnic category 2001 census | 535 |
| 93961000000109 | Italian - ethnic category 2001 census | 33230 |
| 414551003 | Japanese | 340 |
| 92761000000103 | Japanese - ethnic category 2001 census | 6925 |
| 92651000000105 | Kashmiri - ethnic category 2001 census | 1020 |
| 38361009 | Koreans | 1235 |

Table 9: Count of individual ethnicity code use

| | | |
|---|---|---|
| 93981000000100 | Kosovan - ethnic category 2001 census | 1925 |
| 94091000000101 | Kurdish - ethnic category 2001 census | 13850 |
| 94111000000106 | Latin American - ethnic category 2001 census | 7125 |
| 92781000000107 | Malaysian - ethnic category 2001 census | 3760 |
| 94071000000100 | Middle Eastern (excluding Israeli, Iranian and Arab) - ethnic category 2001 census | 16600 |
| 92631000000103 | Mixed Asian - ethnic category 2001 census | 3735 |
| 92721000000106 | Mixed Black - ethnic category 2001 census | 2440 |
| 94021000000104 | Mixed Irish and other White - ethnic category 2001 census | 1525 |
| 315239007 | Mixed ethnic census group | 8585 |
| 976751000000104 | Mixed multiple ethnic groups: White and Asian - England and Wales ethnic category 2011 census | 4930 |
| 977431000000100 | Mixed multiple ethnic groups: White and Asian - Northern Ireland ethnic category 2011 census | 1535 |
| 976731000000106 | Mixed multiple ethnic groups: White and Black African - England and Wales ethnic category 2011 census | 2710 |
| 977411000000108 | Mixed multiple ethnic groups: White and Black African - Northern Ireland ethnic category 2011 census | 1140 |
| 976711000000103 | Mixed multiple ethnic groups: White and Black Caribbean - England and Wales ethnic category 2011 census | 5985 |
| 977391000000108 | Mixed multiple ethnic groups: White and Black Caribbean - Northern Ireland ethnic category 2011 census | 2065 |
| 976771000000108 | Mixed multiple ethnic groups: any other Mixed or multiple ethnic background - England and Wales ethnic category 2011 census | 5830 |
| 977551000000106 | Mixed multiple ethnic groups: any other Mixed or multiple ethnic background - Northern Ireland ethnic category 2011 census | 325 |
| 978051000000102 | Mixed or multiple ethnic groups: any Mixed or multiple ethnic group - Scotland ethnic category 2011 census | 65 |
| 414752008 | Mixed racial group (racial group) | 2910 |
| 94101000000109 | Moroccan - ethnic category 2001 census | 3975 |
| 94121000000100 | Multi-ethnic islands: Mauritian or Seychellois or Maldivian or St Helena - ethnic category 2001 census | 2305 |
| 270464009 | N African Arab/Iranian (NMO) | 1700 |
| 718131000000106 | Nepali | 5570 |
| 186036009 | New Zealand European | 505 |
| 186039002 | New Zealand Maori | 75 |
| 186035008 | New Zealand ethnic groups | 560 |
| 92731000000108 | Nigerian - ethnic category 2001 census | 11965 |
| 94061000000107 | North African - ethnic category 2001 census | 5180 |
| 275594002 | North African Arab (NMO) | 1690 |
| 92561000000109 | Northern Irish - ethnic category 2001 census | 935 |

Table 9: Count of individual ethnicity code use

| | | |
|---|---|---|
| 414978006 | Oriental | 170 |
| 92521000000101 | Other - ethnic category 2001 census | 241880 |
| 186010002 | Other African countries (NMO) | 1310 |
| 186013000 | Other Asian (NMO) | 6430 |
| 92481000000101 | Other Asian background - ethnic category 2001 census | 305220 |
| 315281001 | Other Asian ethnic group | 57510 |
| 92701000000102 | Other Asian or Asian unspecified - ethnic category 2001 census | 15905 |
| 186000006 | Other Black - Black/Asian orig | 625 |
| 185999009 | Other Black - Black/White orig | 1870 |
| 92501000000105 | Other Black background - ethnic category 2001 census | 52135 |
| 92741000000104 | Other Black or Black unspecified - ethnic category 2001 census | 5080 |
| 186017004 | Other European (NMO) | 33265 |
| 186037000 | Other European in New Zealand | 50 |
| 92451000000107 | Other Mixed background - ethnic category 2001 census | 122360 |
| 92621000000100 | Other Mixed or Mixed unspecified - ethnic category 2001 census | 7075 |
| 94041000000106 | Other White European or European unspecified or Mixed European - ethnic category 2001 census | 160190 |
| 92411000000108 | Other White background - ethnic category 2001 census | 1431975 |
| 94051000000109 | Other White or White unspecified - ethnic category 2001 census | 57390 |
| 315279003 | Other black ethnic group | 8495 |
| 976951000000102 | Other ethnic group: Arab - England and Wales ethnic category 2011 census | 4015 |
| 977851000000109 | Other ethnic group: Arab - Northern Ireland ethnic category 2011 census | 150 |
| 978381000000105 | Other ethnic group: Arab, Arab Scottish or Arab British - Scotland ethnic category 2011 census | 280 |
| 976971000000106 | Other ethnic group: any other ethnic group - England and Wales ethnic category 2011 census | 12815 |
| 977871000000100 | Other ethnic group: any other ethnic group - Northern Ireland ethnic category 2011 census | 170 |
| 978401000000105 | Other ethnic group: any other ethnic group - Scotland ethnic category 2011 census | 75 |
| 186005001 | Other ethnic non-mixed (NMO) | 14085 |
| 186021006 | Other ethnic, Asian/White origin | 11335 |
| 186020007 | Other ethnic, Black/White origin | 1060 |
| 186019001 | Other ethnic, mixed origin | 14140 |
| 186022004 | Other ethnic, mixed white origin | 3060 |

Table 9: Count of individual ethnicity code use

| | | |
|---|---|---|
| 186023009 | Other ethnic, other mixed origin | 4105 |
| 94031000000102 | Other mixed White - ethnic category 2001 census | 11610 |
| 94011000000105 | Other republics which made up the former Yugoslavia - ethnic category 2001 census | 1625 |
| 401214002 | Other white British ethnic group | 20850 |
| 186002003 | Pakistani | 103375 |
| 92461000000105 | Pakistani or British Pakistani - ethnic category 2001 census | 540820 |
| 88941000000100 | Polish - ethnic category 2001 census | 215080 |
| 718964009 | Polish Roma | 455 |
| 80208004 | Portuguese | 12365 |
| 92641000000107 | Punjabi - ethnic category 2001 census | 5350 |
| 160531006 | Race: West indian | 80 |
| 718958002 | Roma | 580 |
| 445343003 | Romanian | 86070 |
| 718960000 | Romanian Roma | 975 |
| 86275006 | Samoan | 55 |
| 92541000000108 | Scottish - ethnic category 2001 census | 7655 |
| 88981000000108 | Serbian - ethnic category 2001 census | 1365 |
| 110781000000102 | Sinhalese - ethnic category 2001 census | 310 |
| 36329002 | Slovak | 11000 |
| 718961001 | Slovak Roma | 2500 |
| 92711000000100 | Somali - ethnic category 2001 census | 17665 |
| 186044009 | South East Asian | 1190 |
| 89021000000101 | South and Central American - ethnic category 2001 census | 9300 |
| 86461000000107 | Sri Lankan - ethnic category 2001 census | 27370 |
| 92671000000101 | Tamil - ethnic category 2001 census | 2665 |
| 81560001 | Tongan | 30 |
| 88921000000107 | Traveller - ethnic category 2001 census | 26330 |
| 275601009 | Turkish (NMO) | 2415 |
| 110401000000103 | Turkish - ethnic category 2001 census | 16270 |
| 275602002 | Turkish Cypriot (NMO) | 215 |

Table 9: Count of individual ethnicity code use

| | | |
|---|---|---|
| 93951000000106 | Turkish Cypriot - ethnic category 2001 census | 1415 |
| 270467002 | Turkish/Turkish Cypriot (NMO) | 970 |
| 93921000000101 | Ulster Scots - ethnic category 2001 census | 20 |
| 312859007 | Vietnamese | 2145 |
| 92751000000101 | Vietnamese - ethnic category 2001 census | 4450 |
| 92551000000106 | Welsh - ethnic category 2001 census | 4510 |
| 275592003 | West Indian (NMO) | 360 |
| 977351000000100 | White - Northern Ireland ethnic category 2011 census | 200 |
| 185984009 | White - ethnic group | 920270 |
| 315236000 | White British | 5746445 |
| 494131000000105 | White British - ethnic category 2001 census | 4156730 |
| 315237009 | White Irish | 37325 |
| 494161000000100 | White Irish - ethnic category 2001 census | 25740 |
| 401213008 | White Scottish | 7505 |
| 92441000000109 | White and Asian - ethnic category 2001 census | 92835 |
| 92431000000100 | White and Black African - ethnic category 2001 census | 89460 |
| 92421000000102 | White and Black Caribbean - ethnic category 2001 census | 93915 |
| 976631000000101 | White: English or Welsh or Scottish or Northern Irish or British - England and Wales ethnic category 2011 census | 726825 |
| 976671000000104 | White: Gypsy or Irish Traveller - England and Wales ethnic category 2011 census | 735 |
| 977971000000108 | White: Gypsy or Irish Traveller - Scotland ethnic category 2011 census | 25 |
| 976651000000108 | White: Irish - England and Wales ethnic category 2011 census | 7315 |
| 977951000000104 | White: Irish - Scotland ethnic category 2011 census | 70 |
| 978011000000101 | White: Polish - Scotland ethnic category 2011 census | 2595 |
| 977911000000103 | White: Scottish - Scotland ethnic category 2011 census | 500 |
| 976691000000100 | White: any other White background - England and Wales ethnic category 2011 census | 71540 |
| 978031000000109 | White: any other White ethnic group - Scotland ethnic category 2011 census | 1290 |
| 977931000000106 | White: other British - Scotland ethnic category 2011 census | 2220 |
| 296841000000102 | Yemeni | 590 |
