## Extended Data Figure 1 for "Consistency, completeness and external validity of ethnicity recording in NHS primary care records: a cohort study in 25 million patients’ records at source using OpenSAFELY"

Figure 1: Barplot showing the proportion of 2021 Census and TPP populations (amended to 2021 grouping) per ethnicity grouped into 5 groups (excluding those without a recorded ethnicity). Annotated with percentage point difference between 2021 Census and TPP populations.

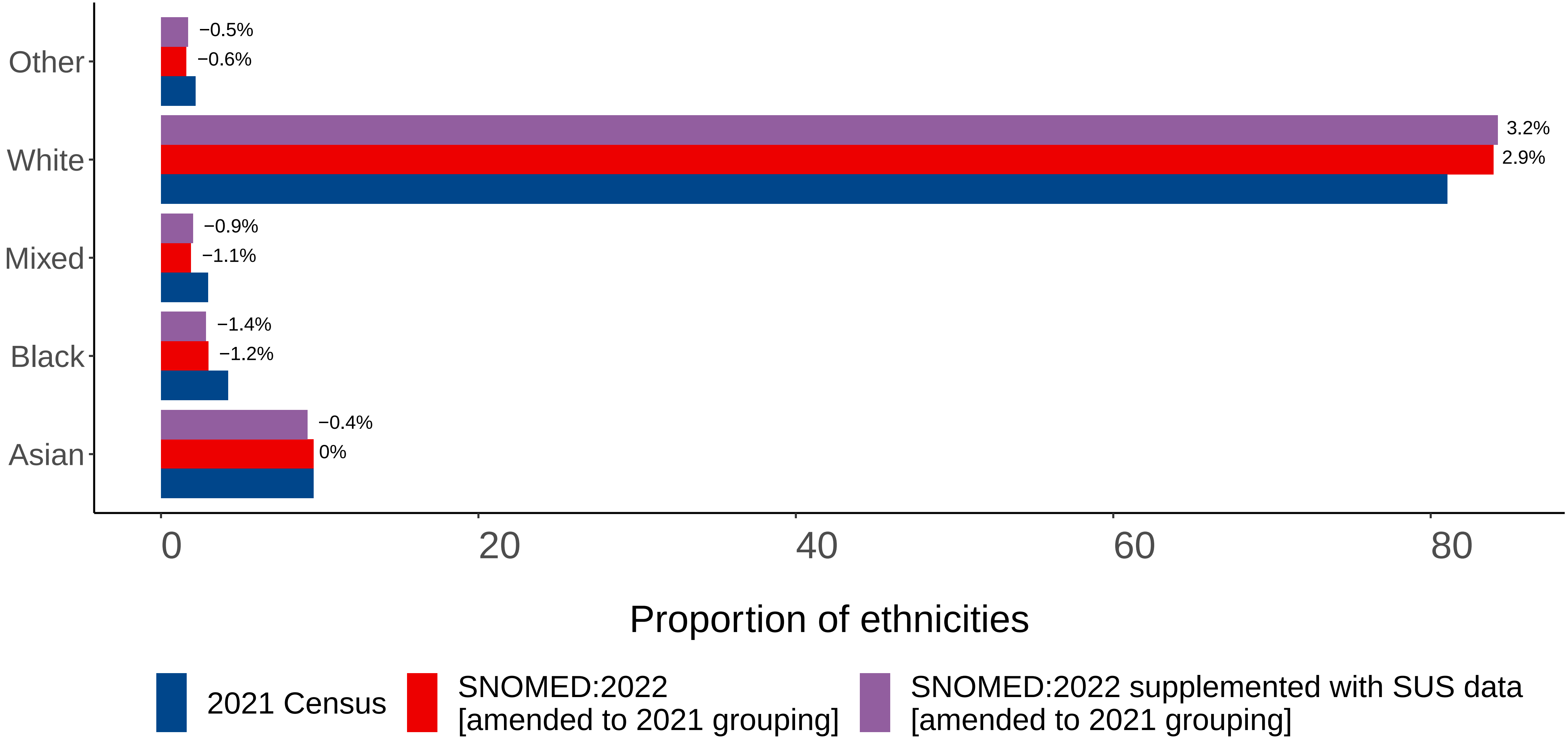
