## Extended Data Figure 2 for "Consistency, completeness and external validity of ethnicity recording in NHS primary care records: a cohort study in 25 million patients’ records at source using OpenSAFELY"

Figure 2: Barplot showing the proportion of 2021 Census and TPP populations (amended to 2021 grouping) per ethnicity grouped into 5 groups per NUTS−1 region (excluding those without a recorded ethnicity). Annotated with percentage point difference between 2021 Census and TPP populations.

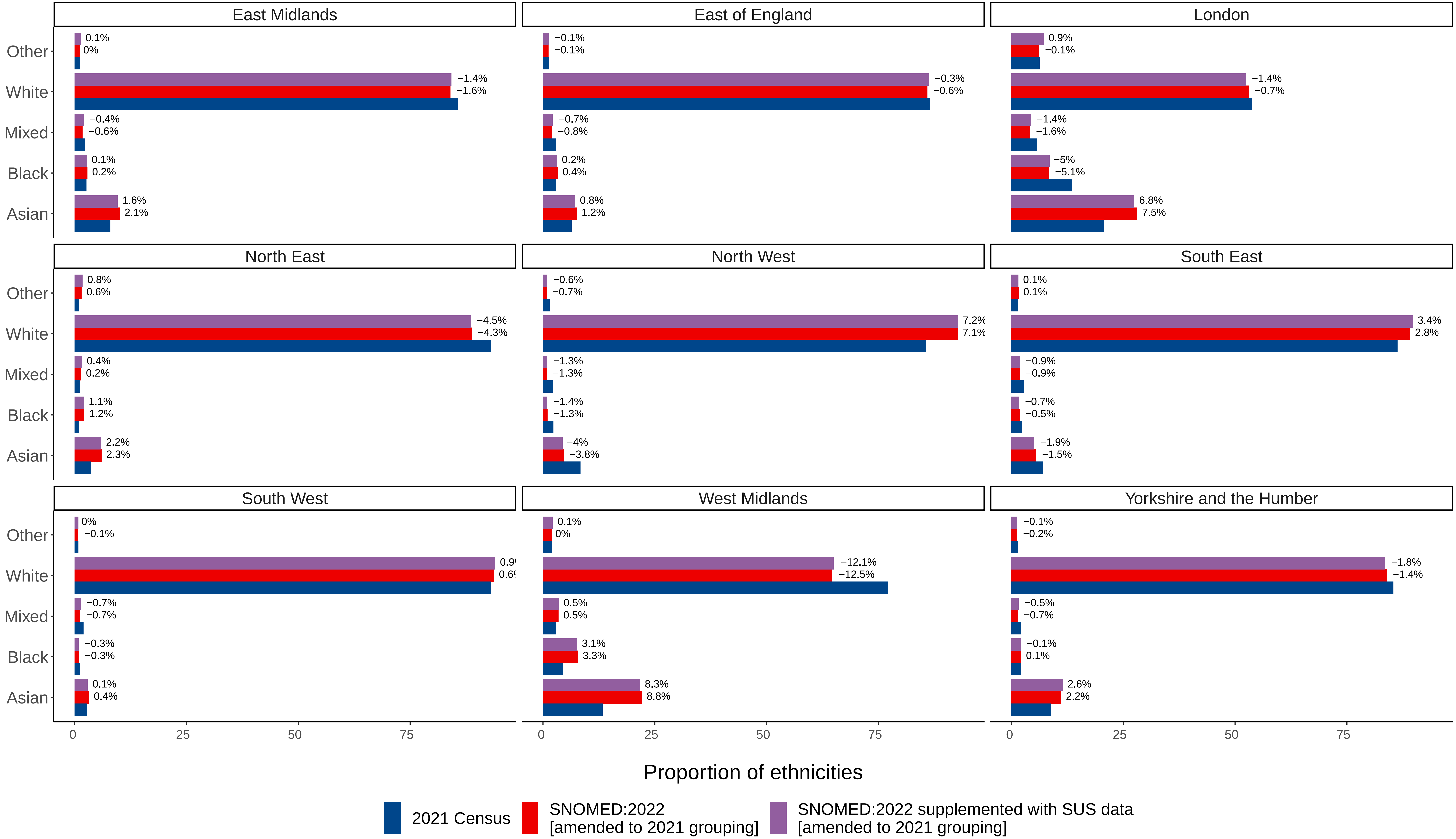
