## Extended Data Figure 2 for "Consistency, completeness and external validity of ethnicity recording in NHS primary care records: a cohort study in 25 million patients’ records at source using OpenSAFELY"

Figure 3: Recording of ethnicity over time for latest and first recorded ethnicity. Unknown dates of recording may be stored as '1900−01−01'

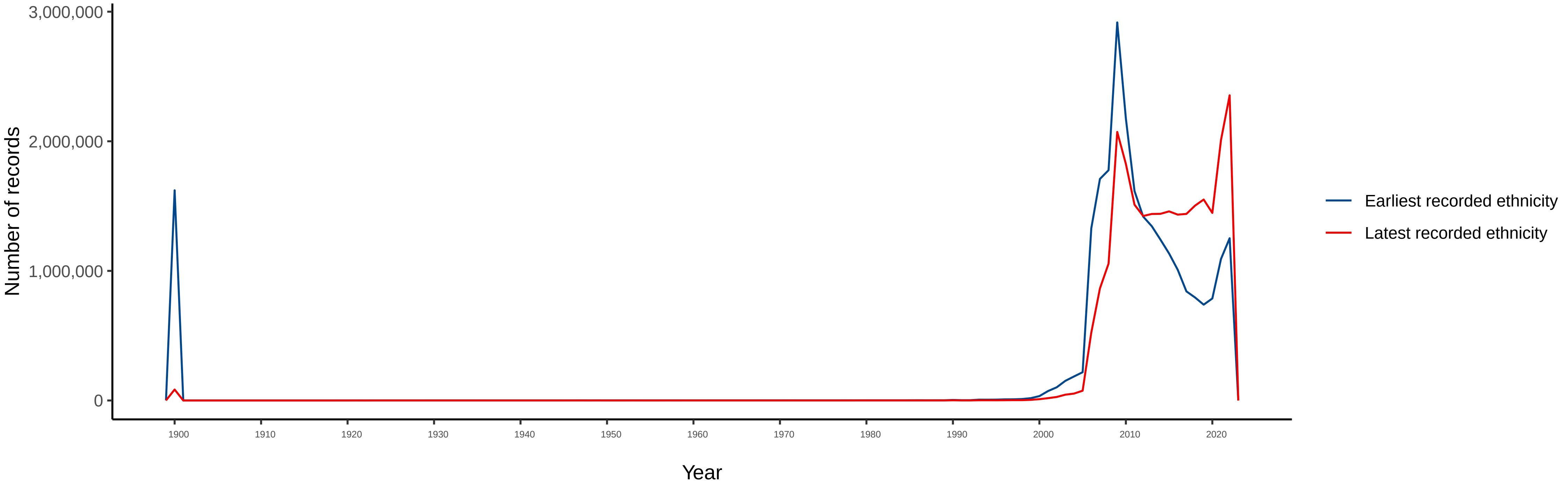
